# Trends of malaria prevalence among individuals from rural communities in three regions with varying transmission intensities in Mainland Tanzania; Data from 2021 - 2023 community cross-sectional surveys

**DOI:** 10.1101/2025.02.13.25322171

**Authors:** Daniel P. Challe, Daniel A. Petro, Filbert Francis, Misago D. Seth, Rashid A. Madebe, Salehe S. Mandai, Rule Budodo, Angelina J. Kisambale, Gervas A. Chacha, Ramadhan Moshi, Ruth B. Mbwambo, Dativa Pereus, Catherine Bakari, Doris Mbata, Beatus Lyimo, Grace K. Kanyankole, Sijenunu Aaron, Daniel Mbwambo, Stella Kajange, Samwel Lazaro, Ntuli Kapologwe, Celine I. Mandara, Vedastus W. Makene, Deus S. Ishengoma

## Abstract

**Background:** Recent reports showed the persistence of malaria transmission and disease burden in rural communities, which have limited the impact of ongoing control and elimination strategies. This study investigated the trends of malaria prevalence among community members from three regions of Mainland Tanzania with varying transmission intensities.

**Methods:** Community surveys were conducted from 2021 to 2023 and involved individuals aged ≥6 months in three regions Kigoma and Ruvuma (with high malaria transmission intensities) and Tanga (moderate transmission). Interviews were conducted using structured questionnaires, to collect anthropometric, clinical, parasitological (testing for malaria using rapid diagnostic tests (RDTs), type of house and socio-economic status (SES) data. Modified Poisson regression was used to identify factors associated with malaria infections and the results were presented as crude (cPR) and adjusted prevalence ratios (aPR).

**Results:** Malaria infections by RDTs were detected in 1,896 (23.2%, n=8,166) individuals, with significant variations across regions and years (22.9% in 2021, 20.6% in 2022, and 26.9% in 2023; p<0.001). The highest prevalence of malaria infections was in Kigoma in 2023 (35.6%) while the lowest was in Tanga in 2022 (10.5%). School children (5 – <15 years) had significantly higher prevalence (38.2% in 2021, 26.2% in 2022, and 34.4% in 2023 (p<0.001) as did males (26.7% in 2021, 25.4% in 2022 and 31.2% in 2023, p<0.001). Higher likelihood of malaria infections was in school children (aPR: 1.94, 95% CI: 1.67 – 2.25, p<0.001), males (aPR=1.24 95%CI: 1.14–1.34, p<0.001), individuals living in traditional houses (aPR=1.14, 95% CI: 1.01 – 1.28, p = 0.037), among individuals with moderate (aPR=1.27, 95% CI: 1.13 – 1.43, p<0.001) or low SES (aPR = 1.39, 95% CI: 1.24 – 1.55, p<0.001), and those with fever at presentation (axillary temperature ≥37.5°C; aPR = 1.34, 95% CI: 1.09 – 1.64, p = 0.005) or fever history within 48 hours before the survey (aPR = 3.55, 95% CI: 3.26–3.87, p<0.001). The likelihood of infections was also higher in Ruvuma (aPR=1.98, 95%CI: 1.77–2.21, p<0.001) and Kigoma (aPR=1.28, 95%CI: 1.15–1.42, p<0.001) regions compared to Tanga. The likelihood of malaria infections was similar among participants based on bed net ownership (aPR: 1.27, 95%CI: 0.80 – 2.01, p = 0.306) or use (aPR: 1.01, 95%CI: 0.64 – 1.50, p=0.920).

**Conclusion:** The study showed spatial and temporal variations of malaria prevalence, with the highest prevalence in 2023 and the lowest in 2022. Groups at higher risk of malaria infections included school children, males, participants with fever, low or moderate SES, and those who lived in traditional houses. Targeted interventions are urgently needed for areas with persistently high transmission and vulnerable groups, particularly in rural communities.

## Background

Malaria remains a significant public health challenge in sub-Saharan Africa (SSA), where it causes substantial morbidity and mortality, particularly among under-fives and pregnant women [1]. In 2023, 263 million malaria cases and 597,000 deaths were reported worldwide, and SSA had 94% of the cases, and 95% of global deaths [2]. Tanzania (4.3%) and the other three SSA countries; Nigeria (30.9%), the Democratic Republic of the Congo (11.3%), and Niger (5.9%) contributed over half of all malaria deaths in 2023 [2]. The majority of malaria cases and deaths reported in SSA are caused by *Plasmodium falciparum*, which is the most virulent malaria parasite species and is responsible for the most severe form of the disease [3–6]. In the World Health Organization’s African Region (WHO Afro-region), other malaria parasite species such as *Plasmodium malariae, Plasmodium ovale,* and *Plasmodium vivax* have also been reported, but their contribution to malaria morbidity and mortality in most of the countries including Tanzania is minimal [7].

Over the past two decades, Tanzania has reported a significant reduction in malaria burden and this has been attributed to the scaled-up interventions as recommended by WHO [8]. Recent reports showed that the number of annual cases has dropped from over 18 million in the 2000s to less than 7 million in 2022, while malaria-related deaths have declined from around 100,000 to less than 7,000 during the same period [9]. Reports have also shown that parasite prevalence in under-fives has declined from 18.1% in 2007 to 7.9% in 2022, despite an increase in parasite prevalence that was reported in 2014 (reaching over 14.0%, above the prevalence of 9.2% in 2012) [10,11]. The burden of malaria within Tanzania has become uneven, with some regions experiencing higher transmission intensities than others [12]. As a result, notable differences exist across regions and districts, with persistently high prevalence (some exceeding 50%) among school children, aged 5 to 16 years in certain areas in north-western and southern regions of Mainland Tanzania [13,14]. These epidemiological changes have been attributed to multiple factors including scaled-up interventions, including the distribution of insecticide-treated bed nets (ITNs) [15], indoor residual spraying (IRS) in some areas [16], and the introduction of artemisinin-based combination therapy (ACTs) [17]. Other factors have also been associated with the current malaria heterogeneity in Tanzania, and they include ecological conditions and vector behaviour [18,19], climate change [20], human behaviours, and the effectiveness of control interventions [21].

Despite the notable progress in reducing malaria prevalence in Tanzania, significant challenges persist, particularly in areas with high and moderate transmission intensities [21,22]. Key obstacles include insecticide resistance in malaria vectors [16], the emergence of drug resistance in some parts of the country [23,24], and parasites with histidine-rich protein 2/3 (*hrp2/3*)) gene deletions that evade the detection by HRP2-based Rapid diagnostic tests (RDT) [25–27]. Other challenges are non-biological challenges, such as reduced funding for malaria control, climate change, and limited intervention coverage worsen the problem [28]. In addition, the temporal dynamics of malaria transmission have been reported in Tanzania, especially among rural communities with limited access to healthcare, but the causes and factors influencing these patterns have remained unexplored and insufficiently understood [29].

Most of the studies on malaria prevalence in rural settings [30–32] have focused on one or more villages from a single geographic area, limiting the ability to track temporal and spatial variations in the prevalence of malaria infections across the country, with special focus on rural areas. Furthermore, these studies often overlook the detailed trends in malaria prevalence in the same communities and rarely account for the temporal and spatial variations as well as changes in the effectiveness of interventions across different regions and transmission settings. This study addresses this gap by investigating the spatial and temporal trends of malaria prevalence in Mainland Tanzania’s rural communities, focusing on regions with high (Kigoma and Ruvuma) and moderate (Tanga) transmission intensities according to the NMCP malaria stratification of 2020. Understanding the temporal trends in malaria prevalence, particularly among rural communities, is crucial for the implementation of evidence-based interventions and strategies as well as targeting specific areas and vulnerable groups to reduce transmission and ultimately eliminate malaria [33]. Therefore, this study aimed to investigate the temporal trends of malaria prevalence among community members (including asymptomatic and symptomatic individuals) living in rural communities located in three different regions of Mainland Tanzania with varying transmission intensities. The findings from this study contribute to the understanding of malaria epidemiology and the recent epidemiological transitions in Tanzania and support the development of more effective and targeted malaria control strategies, with a key focus on areas with persistent transmission and vulnerable groups.

## Methods

### Study design and sites

This study utilised data from community cross-sectional surveys (CSS) that were conducted from 2021 to 2023, covering five villages from three regions of Kigoma, Ruvuma and Tanga in Mainland Tanzania (Figure 1). The surveys were conducted during the peak of malaria transmission in the selected regions in May to June 2021, June 2022 and July to August 2023. These surveys were part of the project on Molecular Surveillance of Malaria in Mainland Tanzania (MSMT), which was conducted in 13 regions in 2021 and 2022, and later expanded to cover all 26 regions of Mainland Tanzania in 2023 as previously reported [34–36]. The selected regions for the CSS had different malaria transmission intensities; with high transmission in Kigoma and Ruvuma and moderate transmission intensities in the Tanga region, based on the 2020 malaria stratification done by the NMCP [26,27,37]. Five villages were covered during the CSS and these included one village each Kigoma (Nyankoronko in Buhigwe district, near the border with Burundi) and Ruvuma (Lundo village in Nyasa District, located on the shores of Lake Nyasa, which borders Malawi) as previously documented [25–27]. The study villages in Tanga region included Magoda, Mamboleo, and Mpapayu (in Muheza district) and these villages were selected because of rich historical data due to their involvement in various malaria studies since 1992 as described elsewhere [30,38–40].

**Figure 1:**
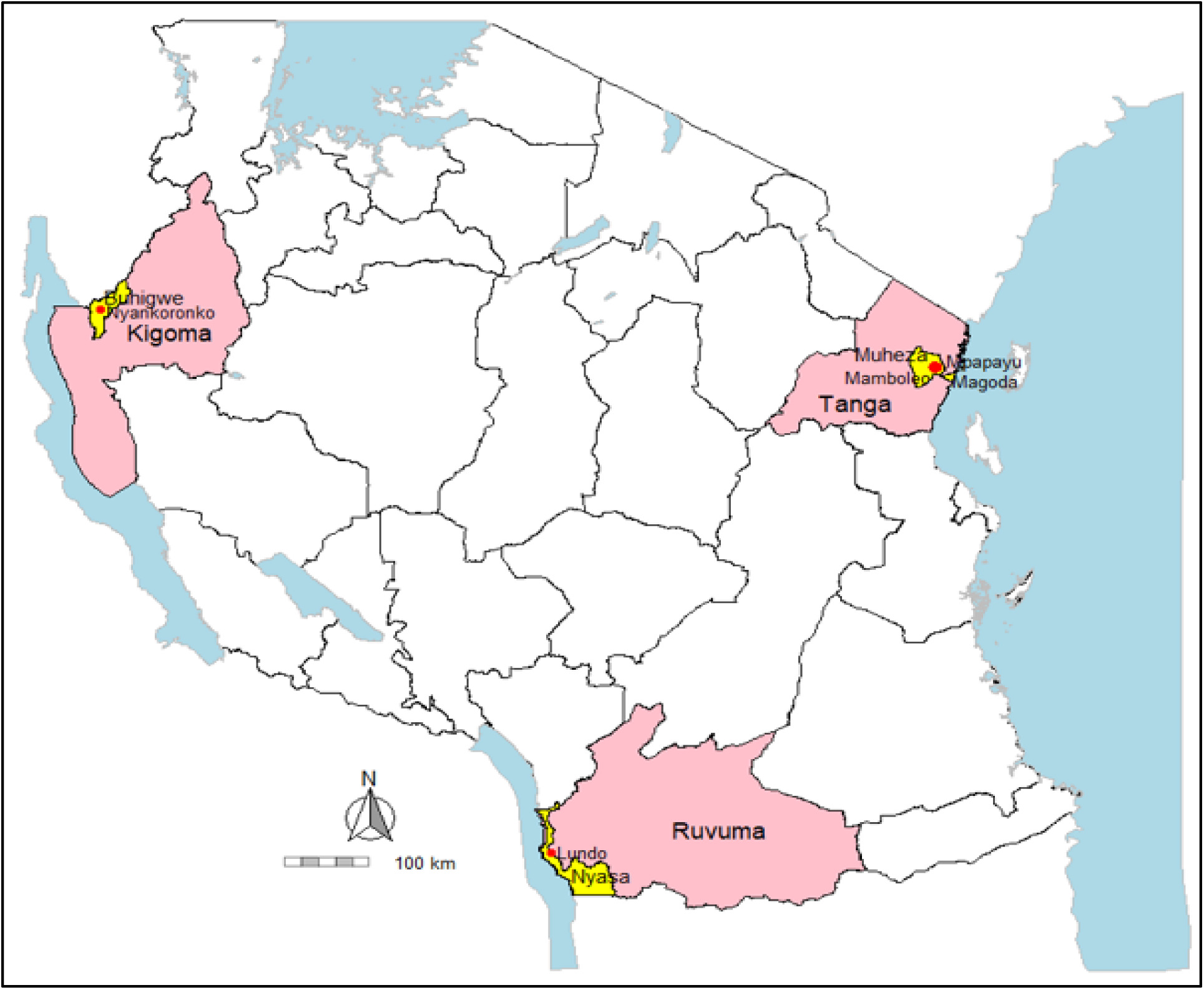
Map of Tanzania showing the sites covered in the CSS with the regions highlighted in pink, districts in yellow and villages represented by the red dots.

### Study population and recruitment of participants

This study targeted and recruited individuals aged 6 months and older residing in the selected villages. The aim was to recruit approximately 30% of registered individuals from each village, based on a census conducted before each CSS. Recruiting about 30% of the target population has been shown to provide a better estimate of the prevalence of malaria infections among community individuals regardless of the symptoms of malaria [31,41]. Eligibility criteria included residence in the study villages, age ≥6 months, and provision of informed consent or assent as previously described [31,41]. Individuals from the villages that were not under the MSMT project or those who did not consent were excluded. Community sensitization teams, under the leadership of village leaders, announced and provided information about the surveys to all community members using loudspeakers and invited willing members to take part in the surveys on the dates allocated to their villages. The project team worked with village leaders to prepare CSS schedules which ensured each village was sufficiently covered based on the number of sub-villages (hamlets) and the total population in the entire village. To enhance accessibility, CSS posts were set up in one of the village hamlets and rotated as needed. Registered members were invited based on hamlets of their residence, and were conveniently recruited, and participation in the CSS was voluntary. Those community members who were unable to attend on their designated day were given another chance to visit the CSS post on another day within the same or nearby villages (for the communities in Tanga which had three villages taking part in the CSS). In each village, the survey took a total of 2 to 5 days depending on the total population in the village, and the team ensured a sufficient number of participants was recruited as previously documented [31,41].

### Data collection procedures

Before conducting the CSS, census surveys were carried out in each village to enumerate and register all households and collect individual’ data in each family, using structured questionnaires. The data captured during the census surveys included demographic, type of house, environmental and socio-economic status (SES) information as previously reported [31,42–44]. Each household in the study villages was properly identified and numbered and all members of the household were given unique identification (IDs) numbers. The house was assessed and allocated to one of the two categories as either traditional or modern houses as previously described [45,46]. A house was considered modern if it was built with bricks and roofed with iron sheets, with or without cement and/or tiles on the floors. A modern house had closed eaves and closed/screened windows. Traditional houses were either made of bricks and thatched roofs, sticks and thatched roofs, mud and thatched roofs, or mud and roofed with iron sheets, and mud or soil floors, open or partially open windows or no windows but with open eaves. Individuals’ and household data including type of houses and SES were updated during annual census surveys, and new members as well as new houses in each of the villages were included in the project database. All data were transferred to a central database at the National Institute for Medical Research (NIMR) in Dar es Salaam.

During the CSS, participants were identified using their unique IDs which were assigned during census surveys and these IDs were generated from the project database before the surveys. Any person who was missing from the list prepared from the project database for the CSS was added to the census list and invited to take part in the CSS. Data collection was done by trained and experienced project staff with the support of trained community health workers (CHWs). The CHWs were from the same villages and were trained and took part in previous surveys, and if needed or in the case of new CHWs, training was done before the surveys. The data were collected using structured questionnaires which are part of the project’s case report forms (CRFs). The CRFs were filled by experienced MSMT project staff and the data collection process included verification of demographic information from the project database and collection of each participant’s anthropometric, parasitological, and clinical data as previously described [31]. Each eligible participant was assigned a unique survey-specific ID and given a registration card with the personal information required for the survey. After registration, participants were interviewed to gather socio-demographic and malaria prevention information. Participants then proceeded to another section for anthropometric measurements (body weight, height, and axillary temperature) which were done by the research team with the help of CHWs. Thereafter, participants proceeded to the laboratory section where finger pricks were performed and blood samples were collected for the detection of malaria parasites using RDTs. The RDTs used in the different CSS included SD Bioline (SD Biosensor, Inc., South Korea) for the 2021 and 2022 surveys while, Abbott Bioline Malaria Ag Pf/Pan (Abbott Diagnostics Korea Inc., Gyeonggi-do, Korea) and Malaria Pf/Pan Ag Rapid Test Cassette (Zhejiang Orient Gene Biotech Co. Ltd, Zhejiang, China) were used in 2023. From the same finger prick, dried blood spots (DBS) on filter papers (Whatman No. 3, GE Healthcare Life Sciences, PA, USA) and blood smears (thin and thick) were collected for further laboratory analysis. Some of the findings of laboratory analyses have been reported elsewhere [35,47]. Participants then went to the final section where clinical assessment was performed by the study clinicians to obtain data on the history of any illness and any treatments taken in the past seven days before the survey. Each participant received a physical examination and a clinical diagnosis was done accordingly. Those who tested positive for malaria were treated according to the national guidelines [17], while participants with other illnesses received appropriate treatment based on their diagnoses [48]. The CSS workflow used in the surveys has been optimized by the MSMT project to ensure smooth and one-directional movement of participants from registration to the clinical section, where they arrive with RDT results. This prevents sending back any participants for laboratory testing in case the clinical section was set up after the laboratory section and participants had to visit clinicians before passing through the laboratory, indicating that two visits to the clinical section would have been warranted as described earlier [31].

### Data management and analysis

All census and CSS data were collected using the MSMT project’s CRFs which were optimized to run on the Open Data Kit (ODK) software installed and run on tablets. The data was collected offline but when the study team got access to the internet, they transferred the data for storage on a central server located at NIMR in Dar es Salaam. Thereafter, the data were downloaded into Excel spreadsheets for cleaning daily, and after completion of the surveys. Additional cleaning was done using Excel and the analysis was performed using STATA version 17 (STATA Corp Inc., TX, USA). Descriptive statistics were used to determine and present the prevalence of malaria for each CSS and the temporal and spatial changes in the prevalence in the study regions. Additional analysis was conducted to investigate the occurrence of malaria in relation to household features, such as house type and SES. The houses were categorized into two types; modern and traditional, as described above and by previous studies [49,50].

The SES for the wealth index was determined using principal component analysis (PCA), as described elsewhere [31,51]. The PCA was used to generate scores from the first component, which were then utilised to categorise households into three wealth quintiles: low, moderate, and high. Members residing in the households falling within the low quintile were considered poor, whereas those in the high quintile were considered rich (or with a higher SES). Modified Poisson regression analysis was used to assess the association between malaria infections and different risk factors including age groups, sex, fever status, year of survey, region, ownership and use of bed nets, type of house, SES and the study village. In this analysis, fever status was defined as the history of fever in the past 48 hours before the survey of fever at presentation with axillary temperature ≥37.5^0^C. Confounding variables were identified and fitted in the multivariate analysis model. The strength of association was presented using both crude (cPRs) and adjusted prevalence ratios (aPRs), with their corresponding 95% confidence intervals (CIs). A chi-square test was used to detect any differences among categorical variables, and a two-sided p-value of ≤0.05 was considered statistically significant for all the analyses.

## Results

### Characteristics of study participants

Overall, 8,166 individuals were enrolled in the study and tested with RDTs. Of the three regions, Tanga had more participants (43.8%, n = 3,575/8,166), followed by Kigoma (30.0%, n = 2,451/8,166) and Ruvuma region (26.2%, n = 2,140/8,166), and the differences in the proportions of participants from the study regions were statistically significant (p<0.001) (Table 1). Most of the participants were adults (aged >15 years) with 47.0% (n = 3,840/8,166), followed by school children (aged 5 - <15 years), who accounted for 38.7% (n = 3,158/8,166) and under-fives who comprised of 14.3% (n = 1,168/8,166) of the participants. Most of the participants were females (58.9%, n = 4,809/8,166) compared to their male counterparts (41.1%, n = 3,357/8,166). The majority of the study participants (85.6%; n = 6,993/8,166) reported owning bed nets, and a large proportion of the participants (85.0%, n = 6,939/8,166) reported having used bed nets the night before the survey. Only 15.0% (n = 1,227/8,166) of the participants did not use bed nets the night before the survey. Of the participants, 30.0% (n = 2,452/8,166) had a history of fever within 48 hours before the study while only 1.9% (157/8,166) had fever at presentation (with axillary temperature ≥37.5°C). The majority of the participants (84.1%, n = 6,343/7,545) lived in traditional houses while only 15.9% (n = 1,202/7,545) lived in modern houses. The distribution of participants based on SES was higher in participants from the low SES category compared to other categories (39.2%, n = 2,962/7,545) (Table 1)

**Table 1:** Characteristics of study participants from the three regions of Kigoma, Ruvuma and Tanga.

| Variable | Total<br>n (%) | Study year |  |  | P-value |
| --- | --- | --- | --- | --- | --- |
|  |  | 2021 n (%) | 2022 n (%) | 2023 n (%) |  |
| Enrolled participants (n) | 8,166 | 2,647 (32.4) | 3,050 (37.4) | 2,469 (30.2) | <0.001 |
| Region |  |  |  |  |  |
| Kigoma | 2,451 (30.0) | 878 (33.2) | 853 (28.0) | 720 (29.2) | <0.001 |
| Ruvuma | 2,140 (26.2) | 741 (28.0) | 905 (29.7) | 494 (20.0) | <0.001 |
| Tanga | 3,575 (43.8) | 1,028 (38.8) | 1,292 (42.3) | 1,255 (50.8) | <0.001 |
| **P-value** | <0.001 | <0.001 | <0.001 | <0.001 |  |
| Age group (Years) |  |  |  |  |  |
| Under-fives | 1,168 (14.3) | 450 (17.0) | 421 (13.8) | 297 (12.0) | <0.001 |
| 5 - <15 | 3,158 (38.7) | 892 (33.7) | 1,260 (41.3) | 1,006 (40.8) | <0.001 |
| ≥15 | 3,840 (47.0) | 1,305 (49.3) | 1,369 (44.9) | 1,166 (47.2) | <0.001 |
| **P-value** | <0.001 | <0.001 | <0.001 | <0.001 |  |
| Sex |  |  |  |  |  |
| Male | 3,357 (41.1) | 1,079 (40.8) | 1,268 (41.6) | 1,010 (40.9) | <0.001 |
| Female | 4,809 (58.9) | 1,568 (59.2) | 1,782 (58.4) | 1,459 (59.1) | <0.001 |
| **P-value** | <0.001 | <0.001 | <0.001 | <0.001 |  |
| Bed net ownership |  |  |  |  |  |
| Yes | 6,993 (85.6) | 2,308 (87.2) | 2,512 (82.4) | 2,173 (88.0) | <0.001 |
| No | 1,173 (14.4) | 339 (12.8) | 538 (17.6) | 296 (12.0) | <0.001 |
| **P-value** | <0.001 | <0.001 | <0.001 | <0.001 |  |
| Bed net use |  |  |  |  |  |
| Yes | 6,939 (85.0) | 2,297 (86.8) | 2,484 (81.4) | 2,158 (87.4) | <0.001 |
| No | 1,227 (15.0) | 350 (13.2) | 566 (18.6) | 311 (12.6) | <0.001 |
| **P-value** | <0.001 | <0.001 | <0.001 | <0.001 |  |
| History of fever in the past 48 hours |  |  |  |  |  |
| Yes | 2,452 (30.0) | 1,295 (48.9) | 729 (23.9) | 428 (17.3) | <0.001 |
| No | 5,714 (70.0) | 1,352 (51.1) | 2,321 (76.1) | 2,041 (82.7) | <0.001 |
| **P-value** | <0.001 | 0.27 | <0.001 | <0.001 |  |
| Fever at presentation (axillary temperature ≥37.5°C) |  |  |  |  |  |
| Yes | 157 (1.9) | 64 (2.4) | 49 (1.6) | 44 (1.8) | 0.13 |
| No | 8,009 (98.1) | 2,583 (97.6) | 3,001 (98.4) | 2,425 (98.2) | <0.001 |
| **P-value** | <0.001 | <0.001 | <0.001 | <0.001 |  |
| House type* |  |  |  |  |  |
| Modern | 1,202 (15.9) | 311 (13.4.) | 539 (18.5) | 352 (15.2) | <0.001 |
| Traditional | 6,343 (84.1) | 2,012 (86.6) | 2,369 (81.5) | 1,962 (84.8) | <0.001 |
| **P-value** | <0.001 | <0.001 | <0.001 | <0.001 |  |
| SES* |  |  |  |  |  |
| High | 2,231 (29.6) | 551 (24.7) | 953 (42.7) | 727 (32.6) | <0.001 |
| Moderate | 2,352 (31.2) | 594 (25.3) | 956 (40.6) | 802 (34.1) | <0.001 |
| Low | 2,962 (39.2) | 1,178 (39.8) | 999 (33.7) | 785 (26.5) | <0.001 |
| **P-value** | <0.001 | <0.001 | 0.13 | 0.50 |  |

SES = socio-economic status, n = number of individuals; \*Missing observation 621

### Prevalence of malaria infections in the study communities

The overall prevalence of malaria infections was 23.2% (n = 1,896/8,166). The prevalence fluctuated over the years, with significant changes in the entire period as shown in Table 2. It decreased from 22.9% (n = 606/2,647) in 2021 to 20.6% (n = 627/3,050) in 2022, followed by a significant rise to 26.9% (n = 663/2,469) in 2023 (p<0.001). The prevalence was significantly higher among school children in each year (p<0.001), with 38.2% (n= 341/892) in 2021, 26.2% (n= 330/1,260) in 2022, and 34.4% (n= 346/1,006) in 2023 compared to other age groups. In each survey, males showed consistently and significantly higher prevalence than females, with the highest prevalence in males observed in 2023 (31.2%, n= 315/1,010) while the lowest was in 2022 (25.4%, n 322/1,268) (p<0.001). Individuals with a history of fever in the past 48 hours had significantly higher prevalence compared to those without a history of fever, and the highest prevalence was reported in 2023 (58.6%, n = 251/428) (p<0.001). For all years (2021 - 2023), the prevalence of malaria was consistently higher among individuals with fever at presentation (axillary temperature ≥37.5°C) compared to those without fever, with statistically significant differences (p<0.001 for 2021 and 2023, and p=0.014 for 2022). The prevalence of malaria infections was significantly higher among participants living in traditional houses compared to those living in modern houses in 2021 (23.8%, n = 478/2,012 vs 18.0%, n = 56/311, p=0.025) and 2023 (27.2%, n = 534/1,962 vs 19.6%, n = 69/352, p=0.003), but no significant difference was observed in 2022 (20.9%, n = 496/2,369 vs 20.6% n = 111/539, p=0.859). The prevalence varied significantly by SES across all years, with the highest observed in the low SES group (25.5%, n = 300/1,178) in 2021, 24.2% (n = 242/999) in 2022, and 34.3% (n = 269/785) in 2023, with significant differences across SES groups (p<0.001). The prevalence of malaria varied by region across the surveys (p<0.001), with the highest prevalence in Ruvuma compared to other regions in 2021 (26.5%, n = 196/741) and in 2022, (29.3%, n = 265/905), while Kigoma had a significantly higher prevalence compared to other regions in 2023 (35.6%, n = 256/720) (p<0.001) (Table 2).

**Table 2:** Prevalence of malaria infections by years and groups.

| Variable | 2021<br>% (n) | 2022<br>% (n) | 2023<br>% (n) | Total<br>% (n) | P-value |
| --- | --- | --- | --- | --- | --- |
| Overall prevalence | 22.9 (606/2,647) | 20.6 (627/3,050) | 26.9 (663/2,469) | 23.2 (1,896/8,166) | <0.001 |
| Age group (Years) |  |  |  |  |  |
| Under-fives | 18.7 (84/450) | 15.9 (67/421) | 18.5 (55/297) | 17.6 (206/1,168) | 0.510 |
| 5 - <15 | 38.2 (341/892) | 26.2 (330/1,260) | 34.4 (346/1,006) | 32.2 (1,017/3,158) | <0.001 |
| $\geq 15$ | 13.9 (181/1,305) | 16.8 (230/1,369) | 22.5 (262/1,166) | 17.5 (673/3,840) | <0.001 |
| P-value | <0.001 | <0.001 | <0.001 | <0.001 |  |
| Sex |  |  |  |  |  |
| Male | 26.7 (288/1,079) | 25.4 (322/1,268) | 31.2 (315/1,010) | 27.6 (925/3,357) | 0.004 |
| Female | 20.3 (318/1,568) | 17.1 (305/1,782) | 23.9 (348/1,459) | 20.2 (971/4,809) | <0.001 |
| P-value | <0.001 | <0.001 | <0.001 | <0.001 |  |
| Bed net ownership |  |  |  |  |  |
| Yes | 22.4 (516/2,308) | 19.8 (498/2,512) | 25.7 (558/2,173) | 22.5 (1,572/6,993) | <0.001 |
| No | 26.6 (90/339) | 24.0 (129/538) | 35.5 (105/296) | 27.6 (324/1,173) | 0.002 |
| P-value | 0.086 | 0.031 | <0.001 | <0.001 |  |
| Bed net use |  |  |  |  |  |
| Yes | 22.4 (515/2,297) | 19.8 (492/2,484) | 25.6 (553/2,158) | 22.5 (1,560/6,939) | <0.001 |
| No | 24.5 (70/286) | 23.7 (112/472) | 34.9 (89/255) | 26.8 (271/1,013) | 0.003 |
| P-value | 0.6131 | 0.053 | 0.002 | 0.003 |  |
| History of fever in the past 48 hours |  |  |  |  |  |
| Yes | 40.1 (519/1,295) | 45.7 (333/729) | 58.6 (251/428) | 45.0 (1,103/2,452) | <0.001 |
| No | 6.4 (87/1,352) | 12.7 (294/2,321) | 20.2 (412/2,041) | 13.9 (793/5,714) | <0.001 |
| P-value | <0.001 | <0.001 | <0.001 | <0.001 |  |
| Fever at presentation (axillary temperature $\geq 37.5^{\circ}\text{C}$) | | | | | |
| Yes | 40.6 (26/64) | 34.7 (17/49) | 50.0 (22/44) | 41.4 (65/157) | 0.32 |
| No | 22.5 (580/2,583) | 20.3 (610/3,001) | 26.4 (641/2,425) | 22.9 (1,831/8,009) | <0.001 |
| P-value | <0.001 | 0.014 | <0.001 | <0.001 |  |
| House type |  |  |  |  |  |
| Modern | 18.0. (56/311) | 20.6 (111/539) | 19.6 (69/352) | 19.4 (236/1,202) | 0.66 |
| Traditional | 23.8 (478/2,012) | 20.9 (496/2,369) | 27.2 (534/1,962) | 24.0 (1,508/6,343) | <0.001 |
| P-value | 0.025 | 0.859 | 0.003 | 0.001 |  |
| SES |  |  |  |  |  |
| High | 16.7 (92/551) | 15.5 (148/953) | 18.2 (132/727) | 16.7 (372/2,231) | 0.36 |
| Moderate | 23.9 (142/594) | 22.7 (217/956) | 25.2 (202/802) | 23.9 (561/2,352) | 0.47 |
| Low | 25.5 (300/1,178) | 24.2 (242/999) | 34.3 (269/785) | 27.4 (811/2,962) | <0.001 |
| P-value | <0.001 | <0.001 | <0.0012 | <0.0014 |  |
| Region |  |  |  |  |  |
| Kigoma | 18.1 (159/878) | 26.5 (226/853) | 35.6 (256/720) | 18.4 (658/3,575) | <0.001 |
| Ruvuma | 26.5 (196/741) | 29.3 (265/905) | 27.5 (136/494) | 27.9 (597/2,140) | 0.43 |
| Tanga | 24.4 (251/1,028) | 10.5 (136/1,292) | 21.6 (271/1,255) | 26.2 (641/2,451) | <0.001 |
| P-value | <0.001 | <0.001 | <0.001 | <0.001 |  |

SES = socio-economic status, n = number of individuals

### Temporal trends of malaria prevalence in the study regions

The prevalence of malaria infections varied significantly among the regions over the three years and Kigoma had a consistent and significant increase in the prevalence from 18.1% (n = 159/878) in 2021 to 26.5% (n = 226/853) in 2022 (p<0.001), and further to 35.6% (n = 256/720) in 2023 (p<0.001). Tanga reported a significant drop in malaria prevalence from 24.4% (n = 251/1,028) in 2021 to 10.5% (n = 136/1,292) in 2022 (p<0.001), followed by a significant increase to 21.6% (n = 271/1,255) in 2023 (p<0.001). Unlike Kigoma and Tanga, the prevalence of malaria in Ruvuma showed slight variations, with a non-significant increase from 26.5% (n = 196/741) in 2021 to 29.3% (n = 265/905) in 2022 (p=0.670), followed by a decrease to 27.5% (n = 136/494) in 2023, but the difference was not statistically significant (p=0.999) (Figure 2A, 2B and 2C).

**Figure 2:**
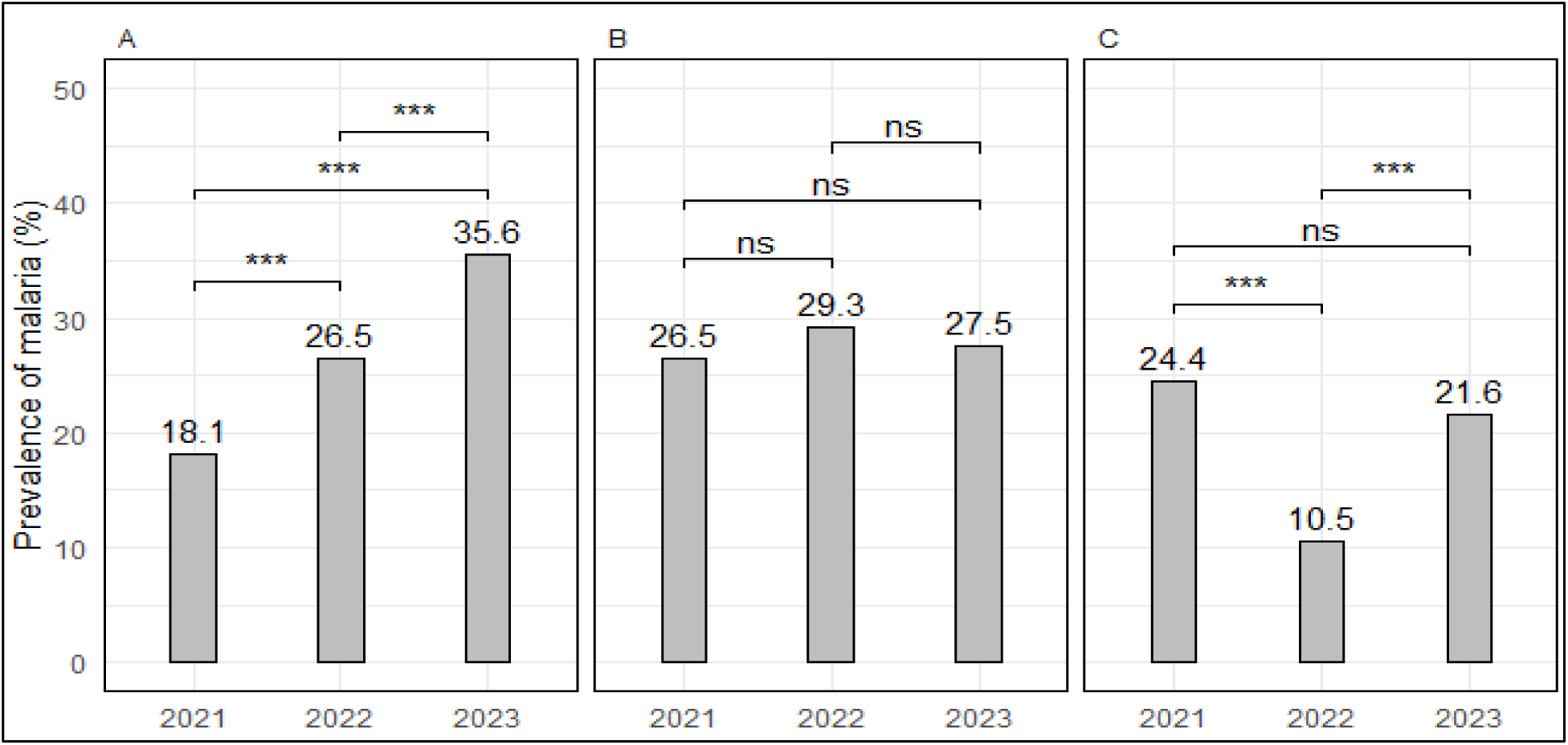
Bargraphs showing temporal patterns of the prevalence of malaria in the three regions Kigoma (A), Ruvuma (B) and Tanga (C) from 2021 to 2023. Note: ***=p<0.001, ns = not statistically significant.

### Age-specific trend patterns of malaria prevalence

Overall, the prevalence of malaria infections was significantly higher in school children across regions, and in all years (p ≤ 0.02) except in Tanga region in 2022 (p = 0.510) (Supplementary Table 1). The prevalence of malaria infections in school children was 38.2% (n = 341/892) in 2021, then it dropped to 26.2% (n = 330/1,260) in 2022 and increased to 34.4% (n = 346/1,006) in 2023 (p<0.001) (Figure 3B). In under-fives, the prevalence was relatively stable, with a slight decrease from 18.7% (n = 84/450) in 2021 to 15.9% (n = 67/421) in 2022 (p=0.33), and an increase to 18.5% (n = 55/297) was observed in 2023, (p=0.42) (Figure 3A). Among adults (aged ≥15 years), the prevalence of malaria infections had a non-significant increase from 13.9% (n = 181/1,305) in 2021 to 16.8% (n = 230/1,369) in 2022 (p = 0.122), and this was followed by a significant increase to 22.5% (n = 262/1,166) in 2023 (p=0.001) as shown in Figure 3C. Based on the prevalence in school children, Kigoma and Ruvuma regions had high transmission with a prevalence >30% in all three years (the prevalence ranged from 33.0% in 2021 to 44.3% in 2023 in Kigoma region). Tanga had high transmission with prevalence of 38.4% in 2021, but the region shifted to moderate transmission in 2022 (prevalence = 10.7%) and 2023 (26.9%). In under-fives, the prevalence was consistently less than 30% in all the surveys and in all three regions (with the lowest of 8.0.3% in Tanga in 2022). For adults (≥15 years), the prevalence in all regions was below 30% in the three years (it ranged from 9.2% in 2021 to 27.7% in the 2023 CSS in the same region of Kigoma (Supplemental Table 1).

**Figure 3:**
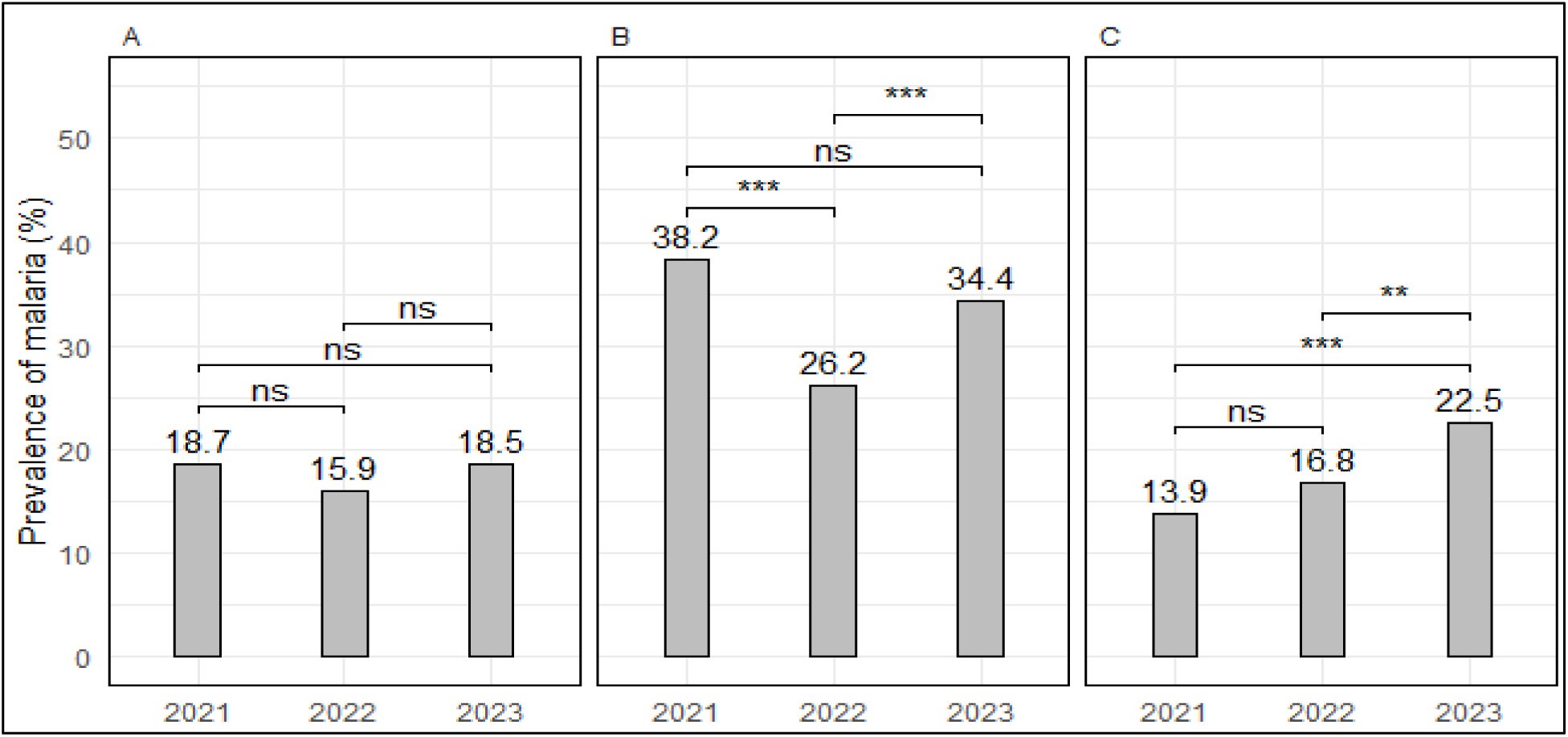
Prevalence of malaria infections among participants of different age groups in the surveys which were undertaken in 2021, 2022 and 2023; A = Under-fives, B = school children (aged 5 - <15 years) and C = adults (≥15 years) The level of significance *P<0.05, **P<0.01, ***P<0.001, ns=not significant

### Sex-specific trends of malaria prevalence

Among participants of different sex, the prevalence of malaria infections decreased between 2021 (26.7%, n = 288/1,079 in males and 20.3%, n = 318/1,568 in females) and 2022 (25.4%, n = 322/1,268 in males and 17.1%, n = 305/1,782 in females) but the differences were not significant (p = 0.999 for males and p = 0.063 for females). However, in 2023, there was a notable increase in malaria prevalence for both males and females, with a significant increase from 25.4% (n = 322/1,268) to 31.2% (n = 315/1,010) among males (p = 0.008) and from 17.1% (n = 305/1,782) to 23.9% (n = 348/1,459) among females (p<0.001) (Figure 4A & 4B). Additionally, in all regions, malaria prevalence among males and females varied significantly but it was significantly and consistently higher in males compared to females (p≤0.01 in all regions and across years) except in Kigoma in 2021 (p = 0.68) and Ruvuma in 2023 (p=0.08) (Supplemental Table 1).

**Figure 4:**
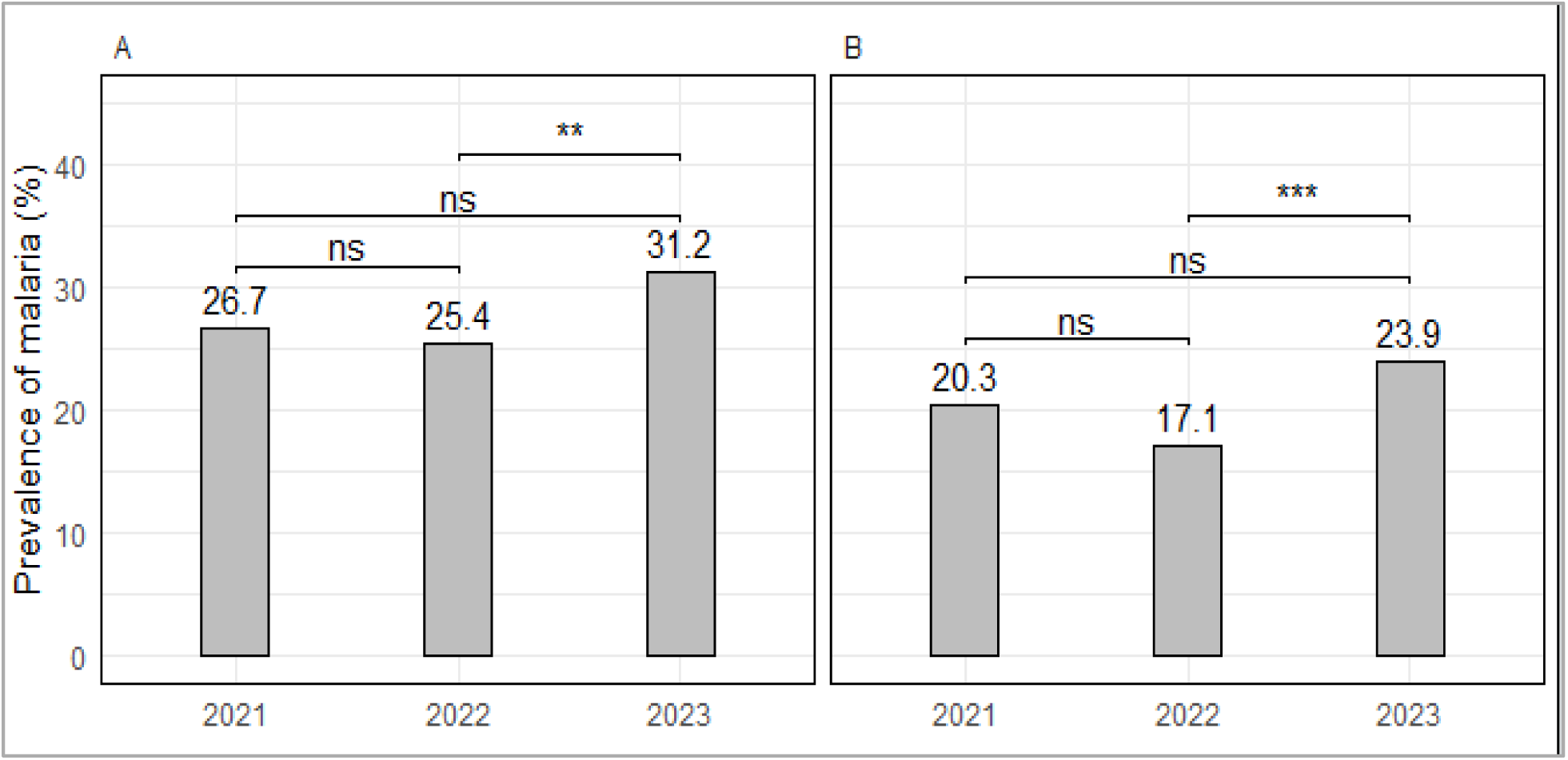
Prevalence of malaria infection among males (A) and females (B) The levels of significance, *P<0.05, **P<0.01, ***P<0.001

### The trends of malaria infections among participants with or without fever

The prevalence of malaria infections in individuals with fever at presentation (axillary temperature ≥37.5°) decreased from 40.6% (n = 26/64) in 2021 to 34.7% (n = 17/49) in 2022 and then increased to 50.0% (n = 22/44) in 2023, however, both decreases and increases were not significantly different (p = 0.654 and p = 0.199, respectively). For individuals without fever at presentation (axillary temperature <37.5°), the prevalence of malaria infections was consistently lower than those reported among individuals who had fever at presentation across all the study years. Among individuals without fever, the prevalence was 22.5% (n = 580/2,583) in 2021, and it decreased to 20.3% (n = 610/3,001) in 2022; followed by an increase to 26.4%, n = 641/2,425 in 2023). However, the differences were not statistically significant for 2021 and 2022 surveys (p = 0.171), while the difference was statistically significant in the 2022 and 2023 surveys (p<0.001) (Figure 5A and B). The prevalence of malaria infections in individuals with a history of fever within 48 hours before the survey, had a consistent and significant increase from 40.1% (n = 519/1,295) in 2021 to 45.7% (n = 333/729) in 2022 (p = 0.049), and further to 58.6% (n = 251/428) in 2023 (p<0.001). In individuals without a history of fever within 48 hours before the survey), malaria prevalence increased significantly from 6.4% (n = 87/1,352) in 2021 to 12.7% (n = 294/2,321) in 2022 (p<0.001), and it further increased significantly to 20.2% (n = 412/2,041) in 2023 (p<0.001) (Figure 5C and D).

**Figure 5:**
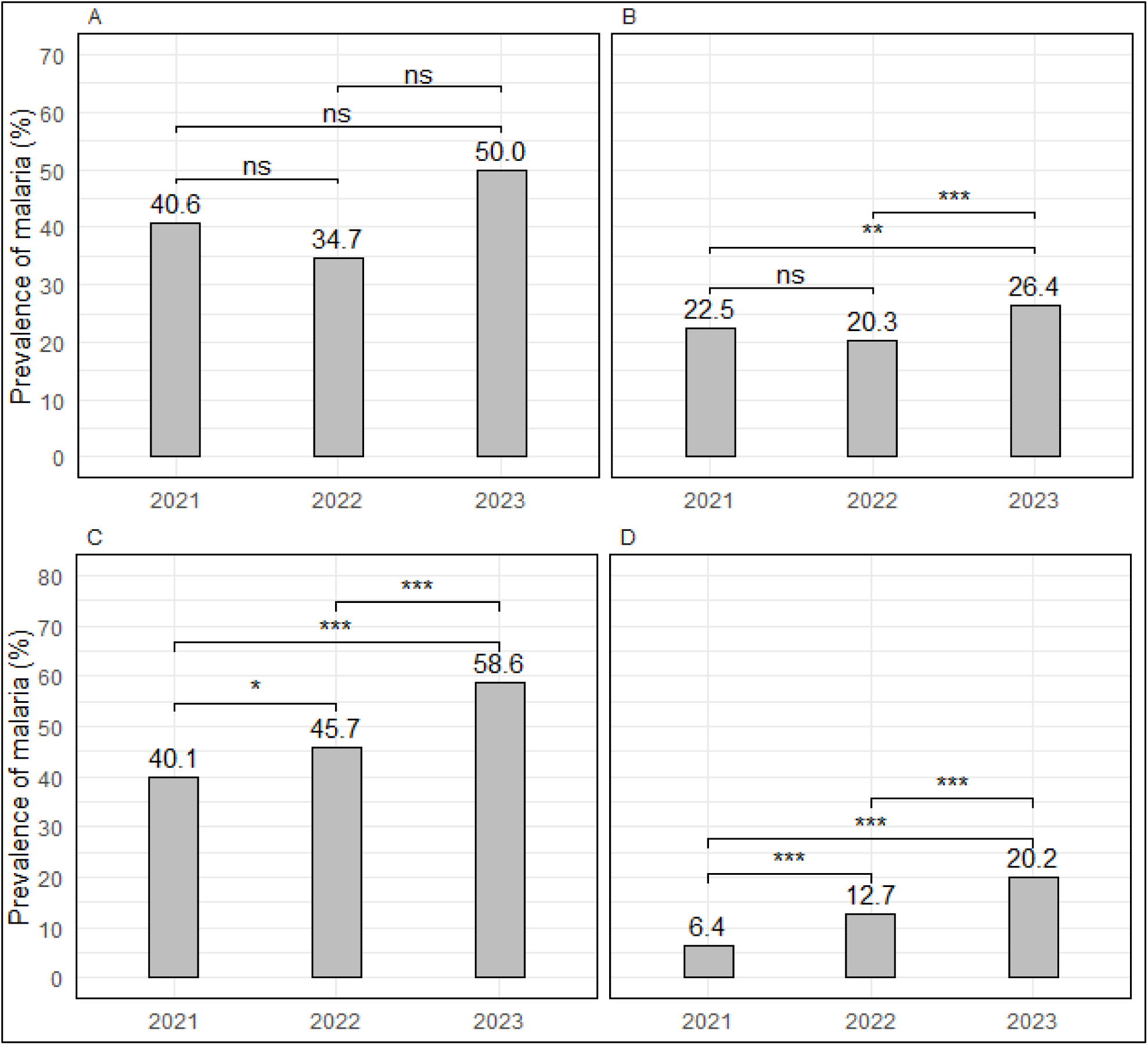
Prevalence of malarial infections among individuals according to their fever status in the surveys which were done in 2021, 2022, and 2023; A = with fever at presentation, B = No fever at presentation, C = history of fever within the past 48 hours, and D = No history of fever in the past 48 hours. Fever at presentation was defined as axillary temperature ≥37.5°C (A) while individuals with axillary temperature ≤37.5°C had no fever. The level of significance is *P<0.05, **P<0.01, ***P<0.001, and ns=not significant.

### Bed net ownership and trends of malaria prevalence

In each region, bed net ownership was 73.0%, 92.7% and 95.3% in 2021; 66.1%, 89.8% and 87.8% in 2022 and 75.7%, 96.0% and 92.0% in 2023 for Kigoma, Ruvuma and Tanga, respectively. While the use of bed nets the night before the survey was reported by 72.6%, 92.4% and 94.8% in 2021; 64.9%, 89.3% and 86.8% in 2022 and 75.3%, 95.3% and 91.2% of the participants for Kigoma, Ruvuma and Tanga, respectively; with no significant differences among the regions (Supplemental Table 2). Among males and females, the use of bed nets was reported majority (≥62.4%) overall in the study years and regions. Based on age groups, the majority reported the use of bed nets was under-fives (90.0%, n = 1,051/1,168) followed by school children (84.8.8%, n = 2,679/3,158) and adults who had the lowest proportion of bed use (83.6%, n = 3,209/3,840) (p<0.001) (Supplemental Table 2). The prevalence of malaria infections among individuals who owned bed nets decreased but not significantly from 22.4% (n = 516/2,308) in 2021 to 19.8% (n = 498/2,512) in 2022 (p = 0.102) and then increased significantly to 25.7% (n = 558/2,173) in 2023 (p<0.001 for 2022 and 2023 comparison). The prevalence of malaria was higher among individuals who did not own bed nets but had a non-significant decline from 26.5% (n = 90/339) in 2021 to 24.0% (n = 129/538) in 2022 (p = 0.998), and a significant increase to 35.5% (n = 105/296) in 2023 (p = 0.001) (Figure 6A & B). For individuals who used bed nets the night before the survey, the prevalence of malaria infections y had a non-significant decrease from 22.4% (n = 515/2,297) in 2021 to 19.8% (n = 492/2,484) in 2022 (p = 088), but this was followed by a significant increase to 25.6% (n = 553/2,158) in 2023 (p<0.001). Among individuals who did not use bed nets, the prevalence of malaria decreased non-significantly from 24.5% (n = 70/286) in 2021 to 23.7% (n = 112/472) in 2022 (p = 0.997), but it increased significantly to 34.9% (n = 89/255) in 2023 (p = 0.005) (Figure 6C & D). The prevalence of malaria across the years was similar in all regions to all participants regardless of bed net use (p ≥ 0.06) while the combined prevalence in all regions for the entire period was higher in individuals who did not use bed nets the night before the survey (p = 0.02) (Supplementary Table 1).

**Figure 6:**
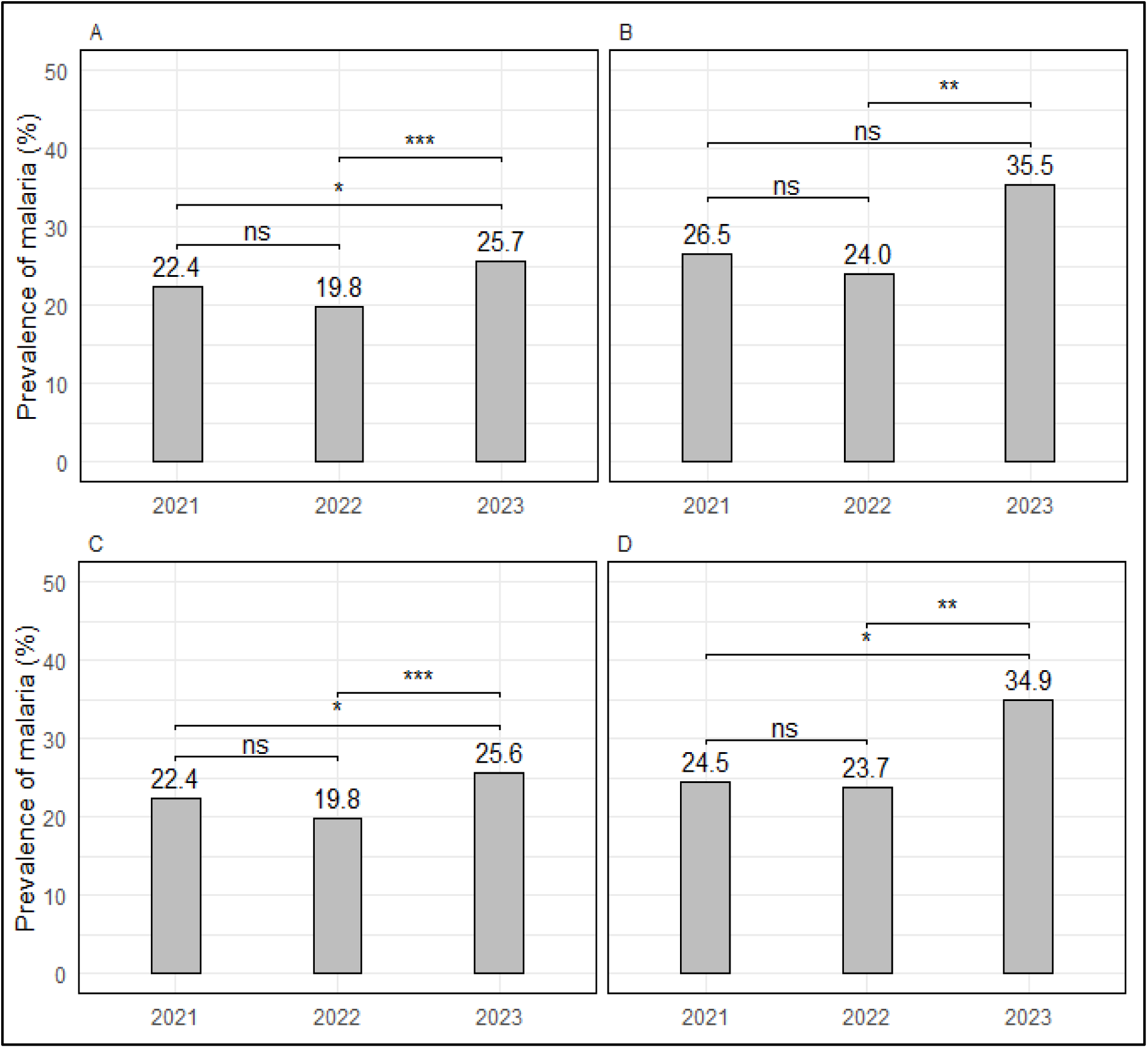
Prevalence of malaria infections among individuals with (A) or without bed nets (B), and in individuals who used (C) or did not use bed nets the night before the survey (D) in 2021, 2022 and 2023. The level of significance *P<0.05, **P<0.01, ***P<0.001, ns=not significant.

### Type of houses, SES and the trends of malaria prevalence

The prevalence of malaria infection among individuals living in modern houses had a non-significant increase from 18.0% (n = 56/311) in 2021 to 20.6% (n = 111/539) in 2022 (p = 0.99), and a non-significant decrease to 19.6% (n = 69/352) in 2023 (p = 0.99). Among individuals living in traditional houses, malaria prevalence decreased non-significantly from 23.8% (n = 478/2,012) in 2021 to 20.9% (n = 496/2,369) in 2022 (p = 0.08), but the prevalence increased significantly to 27.2% (n = 534/1,962) in 2023 (p<0.001) (Figure 7A & B). In the different regions, the prevalence was higher among individuals who lived in traditional houses compared to those from modern houses, with the differences being statistically significant in Tanga (p=0.002) in 2021 and (p=0.002) in 2023 only (Supplementary Table 1).

**Figure 7:**
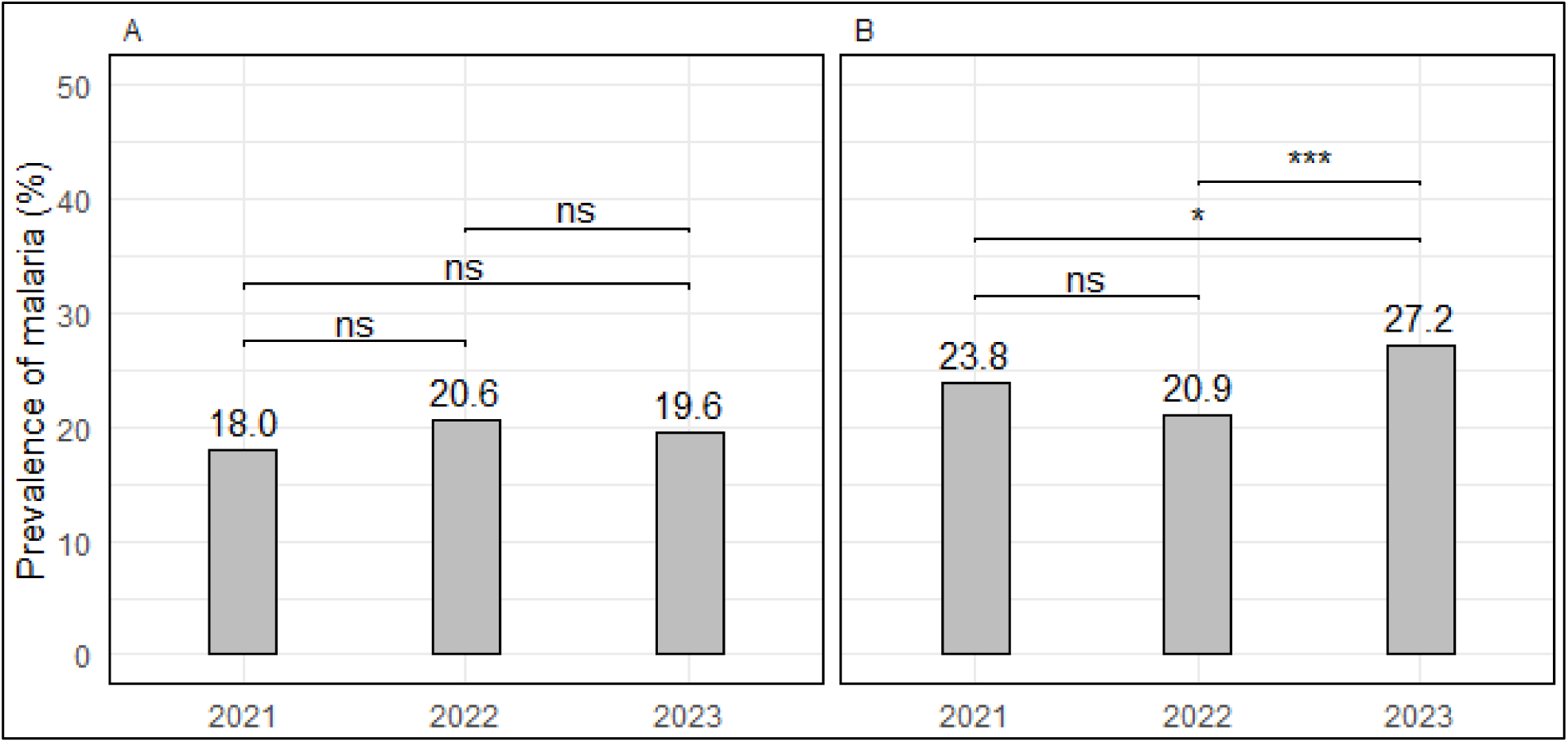
Prevalence of malaria infections among individuals from houses of different types (A = modern and B = traditional houses) in the surveys which were conducted in 2021, 2022 and 2023. The level of significance *P<0.05, **P<0.01, ***P<0.001, ns=not significant.

Malaria prevalence among individuals from households with low SES had a slight but non-significant decrease from 25.5% (n = 300/1,178) in 2021 to 24.2% (n = 242/999) in 2022 (p = 0.999) but increased significantly to 34.3% (n = 269/785) in 2023 (p < 0.001). The prevalence of malaria infections in individuals with moderate SES also had a non-significant decline from 23.9% (n = 142/594) in 2021 to 22.7% (n = 217/956) in 2022 (p = 0.999) while it increased to 25.2% (n = 202/802) in 2023; however, the changes were not statistically significant (p = 0.734). Among individuals with higher SES, the prevalence of malaria infections decreased from 16.7% (n = 92/551) in 2021 to 15.5% (n = 148/953) in 2022, but the decline was statistically not significant (p = 0.995); and this was followed by a non-significant increase to 18.2% (n = 132/727) in 2023 (p = 0.516) (Figure 8A, B & C). The prevalence of malaria infections across the years in Ruvuma and Tanga was significantly higher among participants from households with low SES, except in Tanga region in 2021 when the highest prevalence was among individuals from households with moderate SES (p**≤**0.005 for all comparisons). In Ruvuma region, the prevalence of malaria infections for the three years combined was similar among individuals from households with different levels of SES (p=0.196). For Kigoma region, the prevalence was higher in individuals from families with low SES except in 2022 when the highest prevalence was reported in families with moderate SES but the differences were not statistically significant in all comparisons (in 2022, 2023, and combined analysis for all three years), with exception of 2021 when the prevalence was significantly higher in participants from families with low SES (p=0.014) (Supplementary Table 1).

**Figure 8:**
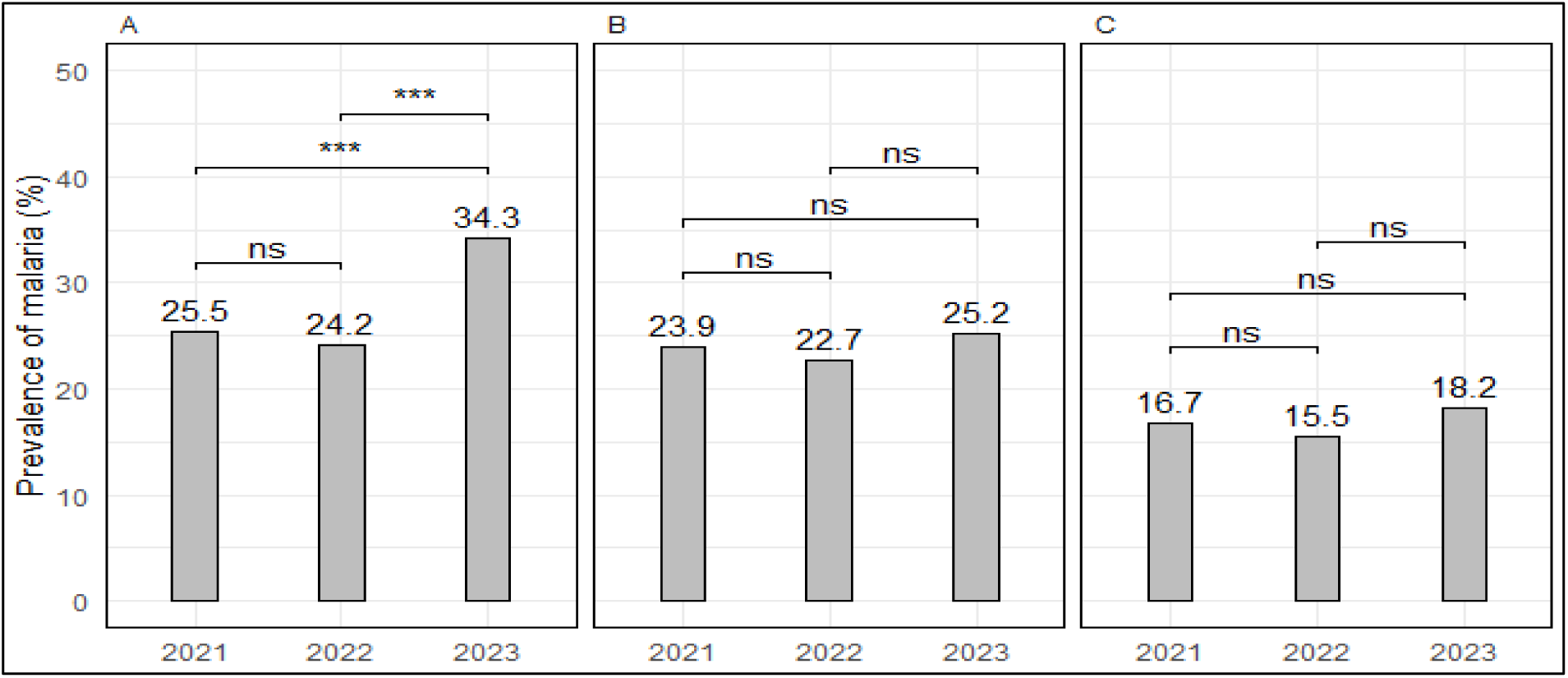
Prevalence of malaria infections among individuals living in households with different levels of SES (A = low, B = moderate and C = high SES) in the surveys which were conducted in 2021, 2022 and 2023. The level of significance *P<0.05, **P<0.01, ***P<0.001, ns=not significant.

### Factors associated with malaria infection in the study area

The likelihood of malaria infections was significantly higher in school children (aged 5 – <15 years (aPR: 1.94, 95% CI: 1.67 – 2.25, p<0.001) and adults (≥15 years; aPR 1.18, 95% CI: 1.01 – 1.38, p = 0.041) when compared to under-fives (Table 3). Males were 1.24 times more likely to have malaria infections (aPR: 1.24, 95% CI: 1.14 – 1.34, p<0.001) compared to females. Among the years, 2023 had nearly two times higher likelihood of malaria infections (aPR: 1.78, 95% CI: 1.60 – 1.98, p<0.001) compared to 2021, and there was a significant difference in the likelihood of malaria infections in 2022 compared to 2021 (aPR: 1.20, 95% CI: 1.08 - 1.33, p = 0.001). After adjusting for different risk factors, the likelihood of malaria infections was similar among all participants regardless of bed net ownership (aPR: 1.25, 95% CI: 0.80 – 1.96, p = 0.324) or use (aPR: 1.01, 95% CI: 0.64 – 1.47, p = 0.893). The likelihood of malaria infections in individuals with fever either at presentation (axillary temperature ≥ 37.5°C) (aPR = 1.34, 95% CI: 1.09 – 1.64, p = 0.005) or history of fever in the past 48 hours before the survey (aPR = 3.55, 95% CI: 3.26–3.87, p<0.001) was significantly higher compared to those who did not have fever. The likelihood of malaria infections was significantly higher in individuals living in traditional houses (aPR=1.14, 95% CI: 1.01 – 1.28, p = 0.037) compared to those living in modern houses. The likelihood of malaria infection was significantly higher among individuals with moderate (aPR=1.27, 95% CI: 1.13 – 1.43, p<0.001) and among individuals with low SES (aPR = 1.39, 95% CI: 1.24 – 1.55, p<0.001) compared to those with higher SES. Among the regions, the likelihood of malaria infections was significantly higher in Ruvuma (aPR = 1.73, 95% CI: 1.57 – 1.91, p<0.001) and Kigoma (aPR = 1.24, 95% CI: 1.12 – 1.37, p<0.001) compared to Tanga region (Table 3).

**Table 3:**
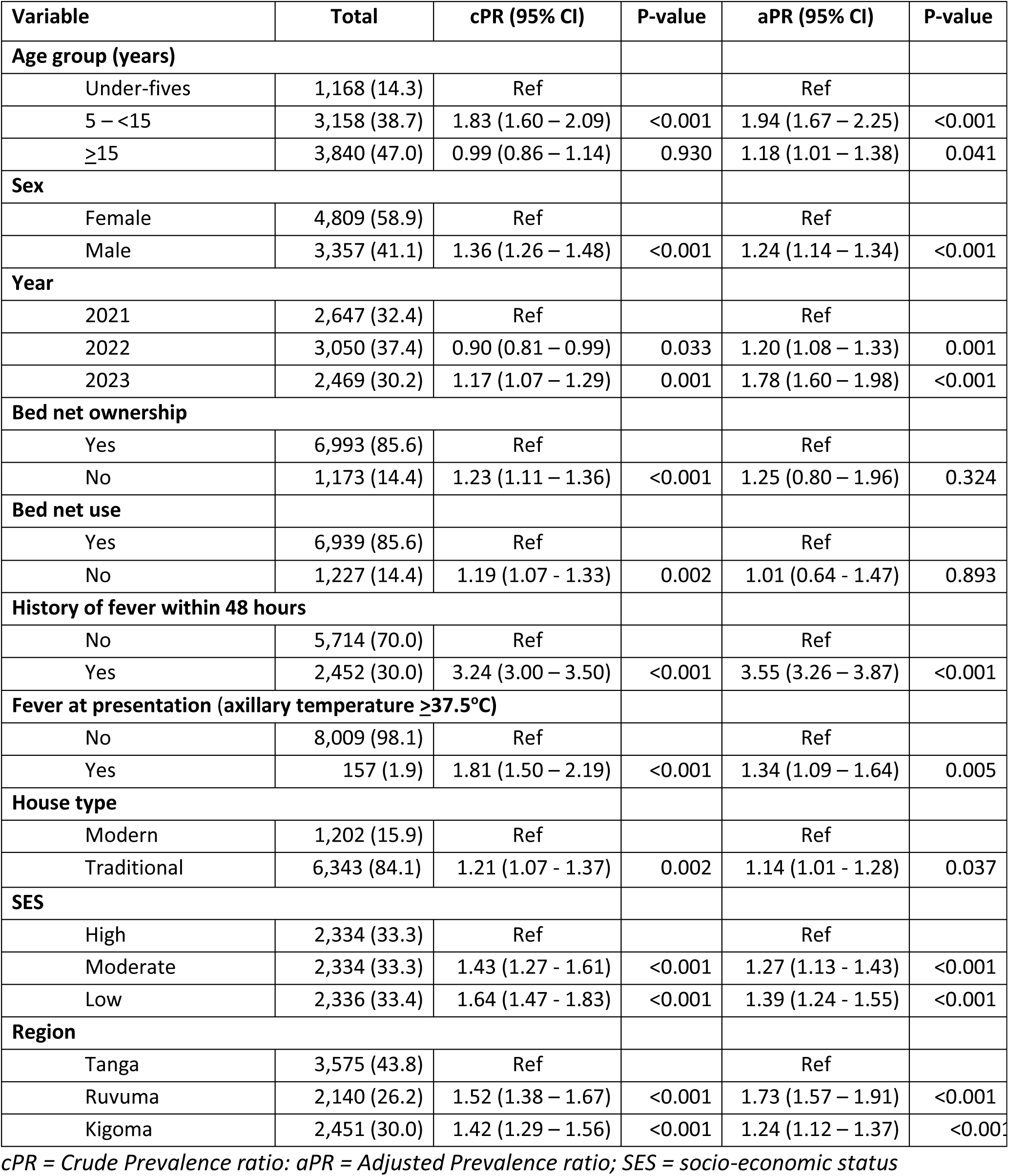
Univariate and multivariate analysis of malaria infections and selected risk factors.

| Variable | Total | cPR (95% CI) | P-value | aPR (95% CI) | P-value |
| --- | --- | --- | --- | --- | --- |
| **Age group (years)** |  |  |  |  |  |
| Under-fives | 1,168 (14.3) | Ref |  | Ref |  |
| 5 – <15 | 3,158 (38.7) | 1.83 (1.60 – 2.09) | <0.001 | 1.94 (1.67 – 2.25) | <0.001 |
| ≥15 | 3,840 (47.0) | 0.99 (0.86 – 1.14) | 0.930 | 1.18 (1.01 – 1.38) | 0.041 |
| **Sex** |  |  |  |  |  |
| Female | 4,809 (58.9) | Ref |  | Ref |  |
| Male | 3,357 (41.1) | 1.36 (1.26 – 1.48) | <0.001 | 1.24 (1.14 – 1.34) | <0.001 |
| **Year** |  |  |  |  |  |
| 2021 | 2,647 (32.4) | Ref |  | Ref |  |
| 2022 | 3,050 (37.4) | 0.90 (0.81 – 0.99) | 0.033 | 1.20 (1.08 – 1.33) | 0.001 |
| 2023 | 2,469 (30.2) | 1.17 (1.07 – 1.29) | 0.001 | 1.78 (1.60 – 1.98) | <0.001 |
| **Bed net ownership** |  |  |  |  |  |
| Yes | 6,993 (85.6) | Ref |  | Ref |  |
| No | 1,173 (14.4) | 1.23 (1.11 – 1.36) | <0.001 | 1.25 (0.80 – 1.96) | 0.324 |
| **Bed net use** |  |  |  |  |  |
| Yes | 6,939 (85.6) | Ref |  | Ref |  |
| No | 1,227 (14.4) | 1.19 (1.07 - 1.33) | 0.002 | 1.01 (0.64 - 1.47) | 0.893 |
| **History of fever within 48 hours** |  |  |  |  |  |
| No | 5,714 (70.0) | Ref |  | Ref |  |
| Yes | 2,452 (30.0) | 3.24 (3.00 – 3.50) | <0.001 | 3.55 (3.26 – 3.87) | <0.001 |
| **Fever at presentation (axillary temperature ≥37.5°C)** |  |  |  |  |  |
| No | 8,009 (98.1) | Ref |  | Ref |  |
| Yes | 157 (1.9) | 1.81 (1.50 – 2.19) | <0.001 | 1.34 (1.09 – 1.64) | 0.005 |
| **House type** |  |  |  |  |  |
| Modern | 1,202 (15.9) | Ref |  | Ref |  |
| Traditional | 6,343 (84.1) | 1.21 (1.07 - 1.37) | 0.002 | 1.14 (1.01 - 1.28) | 0.037 |
| **SES** |  |  |  |  |  |
| High | 2,334 (33.3) | Ref |  | Ref |  |
| Moderate | 2,334 (33.3) | 1.43 (1.27 - 1.61) | <0.001 | 1.27 (1.13 - 1.43) | <0.001 |
| Low | 2,336 (33.4) | 1.64 (1.47 - 1.83) | <0.001 | 1.39 (1.24 - 1.55) | <0.001 |
| **Region** |  |  |  |  |  |
| Tanga | 3,575 (43.8) | Ref |  | Ref |  |
| Ruvuma | 2,140 (26.2) | 1.52 (1.38 – 1.67) | <0.001 | 1.73 (1.57 – 1.91) | <0.001 |
| Kigoma | 2,451 (30.0) | 1.42 (1.29 – 1.56) | <0.001 | 1.24 (1.12 – 1.37) | <0.001 |

cPR = Crude Prevalence ratio; aPR = Adjusted Prevalence ratio; SES = socio-economic status

## Discussion

In most of the malaria endemic countries, more attention and efforts are focused on symptomatic cases that are normally attended at formal health facilities when seeking care for febrile illnesses, while community members mainly including asymptomatic individuals remain largely overlooked. Unlike febrile patients, community members (the majority with asymptomatic infections) are not routinely tested or treated unless they seek care through informal or private healthcare providers. However, such community members carry malaria parasites including gametocytes and contribute to sustained transmission [33]. This study aimed to determine the spatial and temporal trends of malaria prevalence among community members (with or without symptoms of malaria) from three regions of Mainland Tanzania with varying transmission intensities. The study provides insightful data on the trends and patterns of malaria prevalence in these regions and important evidence to support the development of targeted interventions to tackle hotspots and vulnerable groups and enhance malaria control and elimination strategies.

The study showed temporal and spatial variations of the prevalence of malaria infections with relatively high prevalence in all years, and the highest prevalence in 2023 while the lowest was in 2022. Of the three regions, Kigoma reported a consistently higher prevalence of malaria infections, while Tanga had the lowest. Overall, in each region and across the years, the prevalence and risk of malaria infections were significantly higher in males, school children and adults (aged ≥15 years), individuals living in traditional houses, residents of Kigoma and Ruvuma, and those with fever (axillary temperature ≥37.5°C) or a history of fever within 48 hours before the survey. These findings are consistent with those of previous studies which were conducted in the same communities and elsewhere which reported persistence of malaria transmission in some areas and identified some groups with higher prevalence of malaria as vulnerable groups [26]. The observed dynamics of malaria transmission could be due to the availability, utilization, and effectiveness of the current control interventions and other factors that have been shown to impact the spatial and temporal trends, and patterns of malaria transmission in areas of varying endemicity [52]. The findings of this study provide important evidence to support targeting these areas and vulnerable groups with appropriate interventions to reduce the prevalence and disease burden in the affected individuals.

Over the three years, the prevalence of malaria infections was consistently higher in Kigoma with a significant increase each year while the prevalence fluctuated in Ruvuma, with a non-significant increase during the study period. Conversely, Tanga region had a drastic decrease from 2021 to 2022 and a partial rebound in 2023. These changes and patterns of malaria prevalence could be due to various factors such as emerging resistance to insecticides used in bed nets [53,54], increased rainfall [55], inadequate or a lack of impact of malaria control interventions (such as low bed net use in Kigoma, which was at 70.7%), and changes in land use and vegetation cover [56]. Although these were not assessed in this study, changes in land use and vegetation cover together with abnormally high rainfall are the most critical factors linked to creating more breeding sites for mosquitoes and increased transmission intensities within a short period [57]. In Ruvuma region malaria transmission is unimodal [14] and the observed trend could be attributed to seasonal changes that enhance mosquito breeding and subsequently increase malaria transmission intensities[21], potentially emphasizing the need for sustained and timely interventions [30,40]. The changes observed in the communities in Tanga and possibly other regions could also be partially attributed to changes in the amount of rainfall (denoted as excess or deficit of rainfall) which were shown to significantly affect the trends of malaria transmission regardless of the level of control interventions [58]. The regional disparity indicates potential differences in the dynamics of malaria transmission intensities, which could be influenced by factors such as mosquito population density, differences in healthcare-seeking behaviour, availability, access, and use of malaria interventions including vector control and case management as well as health system factors[59–61]. Future studies will need to tease out the different risk factors associated with the persistence of malaria transmission in the regions and other areas of varying endemicity to provide evidence which will allow targeting the key risk factors with area-specific interventions. Addressing such factors will potentially result in more impact within the ongoing efforts to reduce malaria transmission intensities and disease burden and progress to the elimination targets by 2030.

This study reported the lowest prevalence and risk of malaria infections among under-fives while the highest prevalence occurred in school children and adults (≥15 years). The low prevalence of malaria in under-fives could be due to the high coverage of interventions since this age group has been mainly targeted with malaria interventions such as bed nets [62]. Higher prevalence and risk of malaria in school children compared to all other age groups has been widely reported in Tanzania [63] and other countries [64,65] following the epidemiological transition which occurred within the past two decades [8,38]. This has been linked to delays in the development of protective immunity due to delayed exposure to infective bites during childhood caused by unstable malaria transmission in areas that have transitioned from hyperendemic to mesoendemic or hypoendemic transmission [66]. Other factors include an increased risk of exposure to malaria infections, especially in older children [67], and possible gaps in preventive measures targeting this group compared to under-fives and pregnant women [68]. School children are also more active outdoors, especially late in the evening, increasing their likelihood of mosquito bites [69], and may be less diligent in using bed nets or adhering to other preventive practices leading to an increased risk of malaria infections [70]. The observed high prevalence of malaria infection among adults (≥15 years) suggests an increase in the exposure or decreased effectiveness of malaria interventions as reported in other studies [71,72]. While adults typically have higher levels of acquired immunity due to repeated exposure [73], the prevalence of malaria which was higher than that of under-fives could be influenced by factors such as behavioural changes [74] and climatic changes [75]. Additionally, limited access to and use of interventions, economic conditions, and occupational exposure particularly for those working in agriculture or outdoor settings could further increase the risk of malaria infection, especially in rural or economically disadvantaged areas [22] such as the villages covered in this study [76,77]. However, previous studies conducted in these communities [31,41] and other areas [78,79] showed that adults (≥15 years) had a lower prevalence of malaria infections compared to school children and under-fives. Thus, the difference between this study and others cited above will need to be critically assessed in future studies to provide a clear picture and reports of the most vulnerable groups in the communities.

The assessment of sex-specific trends of malaria prevalence indicated a decline in malaria prevalence in 2022, followed by a notable increase in 2023 and males had a higher prevalence compared to females. Higher prevalence in males particularly among older boys and young men and the pattern reported in this study is similar to what has been reported in these communities [80] and elsewhere [81]. The high prevalence and risk of infections among older males could be due to greater exposure that may be linked to occupational and outdoor activities such as farming, fishing, and hunting which increase their exposure to mosquito bites [82]. Additionally, cultural and social norms may influence healthcare-seeking behaviour, with males being potentially less likely to seek early diagnosis and treatment compared to females, leading to a higher prevalence of malaria infections [83]. In contrast, low prevalence among females could likely be due to the more consistent use of malaria interventions like bed nets and chemoprevention (eg. intermittent preventive treatment in pregnancy), as well as high access to healthcare services related to reproductive health and childcare, facilitating early malaria detection and treatment [84,85]. Moreover, previous studies showed that there might be sex-specific immune responses to malaria, with females possibly developing stronger immune responses due to hormonal differences [86], though this area requires further investigation.

The prevalence of malaria infections among individuals with a history of fever within two days before the study or fever at presentation (axillary temperature ≥37.5^0^C) had a similar trend to the overall prevalence of malaria infections, with a decline between 2021 and 2022, followed by an increased prevalence in 2023. Over the three years and in the three regions, individuals with fever had a high risk and correspondingly higher prevalence of malaria infections and this was similar to what has been widely reported by other studies [87]. The high prevalence and risk of malaria infections among febrile participants are believed to be due to limited access to essential services for the prevention, diagnosis and treatment, social and economic disparity, behaviour and roles which increase exposure to infective bites and faster transmission cycle. However, malaria prevalence among individuals with no history of fever 48 hours before the survey increased overtime indicating an increase in the burden of asymptomatic or subclinical infections which pose challenges in malaria control as these individuals can serve as reservoirs of infections and therefore perpetuate malaria transmission in the communities [88,89]. The reasons for this pattern and trends of malaria among individuals with or without fever could be multifactorial, including changes in vector control measures, treatment efficacy, or differences in climatic conditions affecting mosquito populations as reported by other studies [90–92]. This study highlights the relevance of recent febrile episodes as a key symptom to consider in malaria surveillance [93,94] and early detection strategies [95].

The findings showed that individuals who owned and /or used bed nets experienced a lower prevalence of malaria infections with a decrease in prevalence in 2022 highlighting the importance of bed nets as a crucial intervention in malaria control [96,97]. Individuals who did not own or use bed nets had slightly increased likelihood and prevalence of malaria infection emphasising the protective role of bed nets in preventing mosquito bites [70] However, the observed increase in malaria prevalence in 2023, despite high bed net ownership and usage, suggests that other factors such as climatic changes, behavioural factors, bed net condition, and insecticide resistance could have influenced malaria transmission in these communities [98]. The fluctuating trends of malaria prevalence despite high levels of bednet usage in some of the communities (bed net use was ≥87% in Ruvuma and Tanga, but it was lower in Kigoma at ≥65%), indicates the need for continuous efforts in malaria control including regular bed net distribution, public health education on proper usage and addressing potential challenges like insecticide resistance. Furthermore, the increase in malaria prevalence in 2023 suggests that further investigations are needed to determine other factors that may have contributed to the observed trends of malaria prevalence reported in the study communities. Thus, public health initiatives should further promote and facilitate the distribution and use of bed nets to reduce malaria infection.

Individuals living in traditional houses exhibited a significantly higher prevalence and likelihood of malaria infections than those who were living in modern houses. These findings are similar to what was reported in the study that was conducted in Uganda, which showed that there was a significant positive correlation between the prevalence of malaria infections and parasitemia, and the proportion of individuals from traditional houses [99]. While the prevalence of malaria infections among individuals living in modern houses fluctuated slightly, it remained relatively stable. Conversely, individuals living in traditional houses experienced a significant increase in 2023 which was similar to the pattern reported in other groups. Since the majority of the study participants (over 84%) were living in traditional houses, the higher prevalence of malaria in this group suggests that house improvement and poverty reduction strategies are urgently needed. Such interventions might have a higher impact in reducing the disease burden as reported by other studies [49,99,100], and potentially contribute to attaining malaria elimination[101–103]. Previous studies which were conducted in some or all of these communities [31,41], showed that there were no significant differences in the proportion of individuals from families with different levels of SES, but individuals from families with low SES had a higher risk of malaria infections [31,41]. This study showed that the prevalence of malaria infections was significantly higher among individuals with low or moderate SES in Ruvuma and Tanga regions only, while no differences were reported in Kigoma except in 2021. The reasons for these regional disparities are not known and future studies will need to investigate the factors that are associated with a lack of differences in the prevalence of malaria infection among individuals living in households with varying levels of SES in Kigoma region.

This study had some limitations which could affect generalisation of the reported findings. The study covered only three out of the 26 regions in Mainland Tanzania. Two of the regions (Kigoma and Ruvuma) were located in the high transmission stratum while one region (Tanga) was located in the moderate transmission stratum, and no region was covered from low and very low transmission strata. The findings may not be generalisable to other malaria-endemic regions in Mainland Tanzania or beyond due to variations in transmission dynamics and intervention coverage. Detection and confirmation were done by RDTs which may have missed some individuals with subpatent infections or low-density infections as recently reported in some communities in Tanzania [104]. In addition, RDTs especially those based on the detection of HRP2 antigens are associated with high rates of false positives due to the persistence of the antigens after clearance of parasites by treatment or acquired immunity. Using only RDTs may have potentially underestimated or overestimated the prevalence of malaria infections. During the surveys, sampling of study participants was done conveniently by recruiting individuals who volunteered to take part in the study from the target communities with no prior selection of participants. This may have influenced participation in the study, as individuals could have been drawn by the free services which were provided by the project, including medications and consultation which were offered during the survey. This could have led to over-representation or under-representation of some groups potentially affecting the generalizability of the findings. In addition, the data were collected during cross-section surveys which are unlikely to capture temporal variations which are associated with seasonal variations of malaria that may be captured using longitudinal sampling. However, the results of this study are similar to what have been reported by previous studies suggesting that there was a minimal degree of selection bias and the findings may reflect the true prevalence of malaria infections in the communities.

### Conclusion

The study showed temporal variations of malaria prevalence with the highest prevalence in 2023 (26.9%) while the lowest was in 2022 (20.6%) and groups at higher risk of infections included school children, males, participants with fever, those who lived in traditional houses, the years 2022 and 2023, and those living in Ruvuma and Kigoma regions. Targeted interventions are urgently needed for areas with persistently high transmission, and targeting vulnerable groups, particularly in rural communities.

## Data Availability

The dataset of this study is available upon a reasonable request from the corresponding author, with institutional approval by NIMR and the signing of a data transfer agreement (DTA) between NIMR and the recipient.

## Declarations

### Ethical clearance

The ethical approval to conduct this study was obtained from the Medical Research Coordinating Committee of the National Institute for Medical Research (NIMR) in Tanzania. Permission to conduct the study in the villages was provided by the President’s Office, Regional Administration and Local Government Authority, and health authorities of Tanga, Kigoma, and Ruvuma regions, districts, and villages authorities. Informed consent/assent was sought and obtained before conducting the study, from each participant or parents/legal guardians of children. Permission to publish this paper was sought and given by the Director General of the NIMR.

### Consent for publication

Not applicable.

### Competing interests

All authors declare that they have no competing interests.

### Funding

This work was supported in full by the Bill & Melinda Gates Foundation [grant number 002202]. Under the grant conditions of the Foundation, a Creative Commons Attribution 4.0 Generic License has already been assigned to the Author Accepted Manuscript version that might arise from this submission.

### Author’s contributions

DPC, VWM and DSI conceptualised and designed the study and supervised data collection. All authors participated in data collection under the supervision of DSI, and VWM. DPC, FF and DAP managed and analysed data. DPC and DSI wrote the manuscript with the support of VWM and FF, DAP, MDS, CIM, RAM, SSM, RB, AJK, GAC, RM, RBM, DP, CB, DM, BL, GKK, SA, DM, KK, SL and NK and made critical revisions to the initial draft and the final version. All authors read and approved the final version of the manuscript.

## Acknowledgment

The authors wish to thank all study participants for their readiness to participate and take part in this study. They highly appreciate the support of village leaders, districts, and regional medical authorities to the study team during the entire study period. The study was executed by a committed study team including Ezekiel Malecela, Muhidin Kassim, Juma Tupa, Gineson Nkya, Neema Barua, Francis Chambo, Sawaya Msangi, Ally Idrisa, Kusa Mchaina, Christian Msokame, Rogers Msangi, Salome Simba, Mwanaid Mtui, Rehema Mtibusa, Ambele Lyatnga, August Nyaki, Tumaini Kamna, Juma Akida, Athanas Mhina, Salome Simba, Tilaus Gustav, Godbless Msaki, Hatibu Athuman, Anangisye Malabeja, Emmanuel Kessy, George Gesase, Oswald Oscar, Gasper Lugela, Rashid Mtumba, Richard Makono and Ildephonce Mathias. The finance, administrative, and logistic support team at NIMR: Christopher Masaka, Millen Meena, Beatrice Mwampeta, Gracia Sanga, Neema Manumbu, Halfan Mwanga, Arison Ekoni, Twalipo Mponzi, Pendael Nasary, Denis Byakuzana, Alfred Sezary, Emmanuel Mnzava, John Samwel, Daud Mjema, Seth Nguhu, Thomas Semdoe, Sadiki Yusuph, Alex Mwakibinga, Rodrick Ulomi and Andrea Kimboi. Technical and logistics support from the Bill and Melinda Gates Foundation team is highly appreciated.

## Abbreviations and acronyms

ACTs: Artemisinin-based combination therapy
CI: Confidence interval
CSS: Cross-sectional survey
DBS: Dried blood spots
DTA: Data transfer agreement
IPTi: Intermittent preventive treatment in infants
IRS: Indoor residual spraying
ITNs: Insecticide-treated bed nets
IQR: Interquartile range
RDT: Rapid Diagnostic Test
MSMT: Molecular Surveillance of Malaria in Mainland Tanzania
NIMR: National Institute for Medical Research
NMCP: National Malaria Control Program
ODK: Open Data Kit software
PO-RALG: President’s Office, Regional Administration and Local Government
PR: Prevalence Ratio
SSA: Sub-Saharan Africa

---

**Supplementary Table 1:**
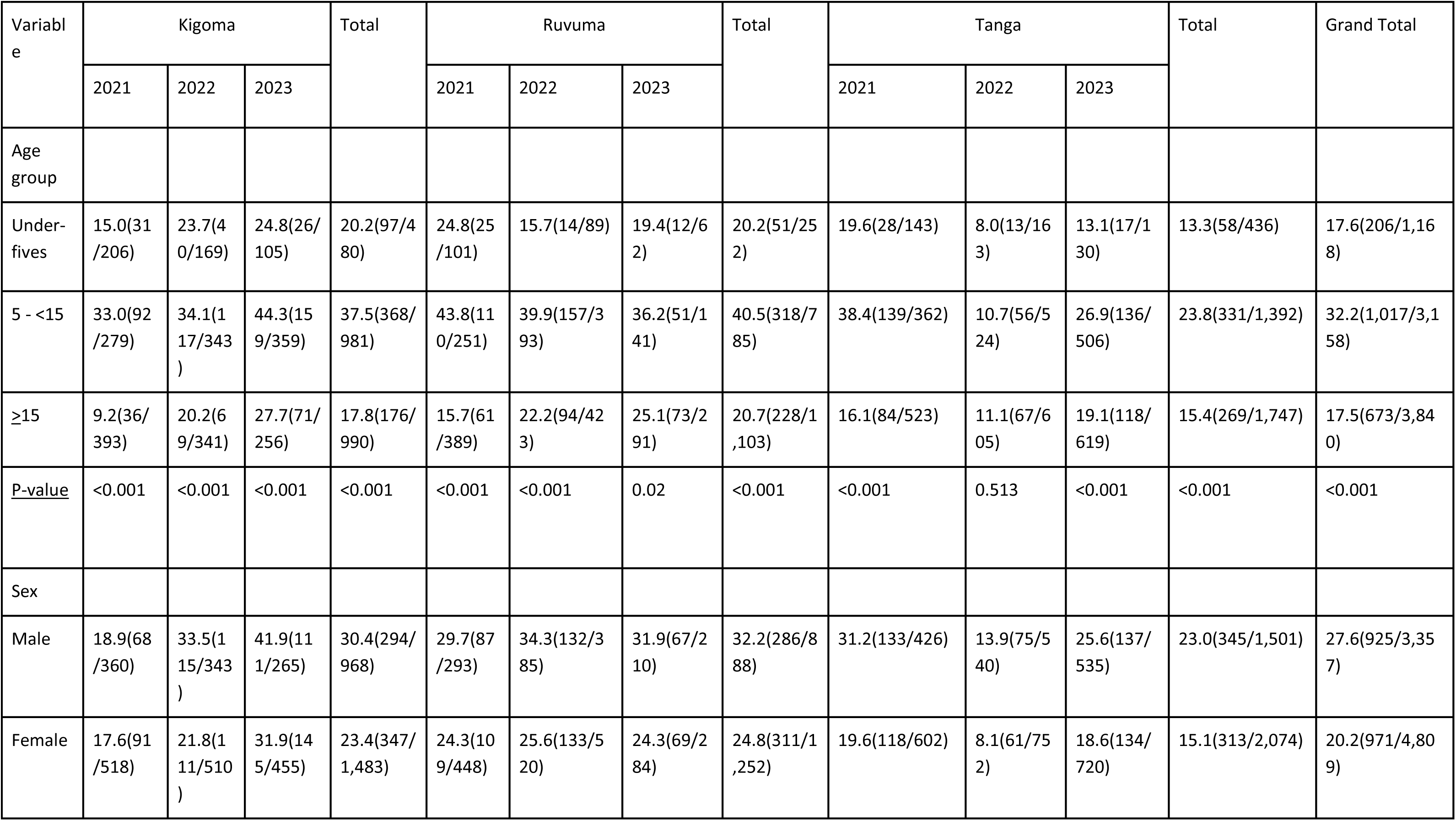
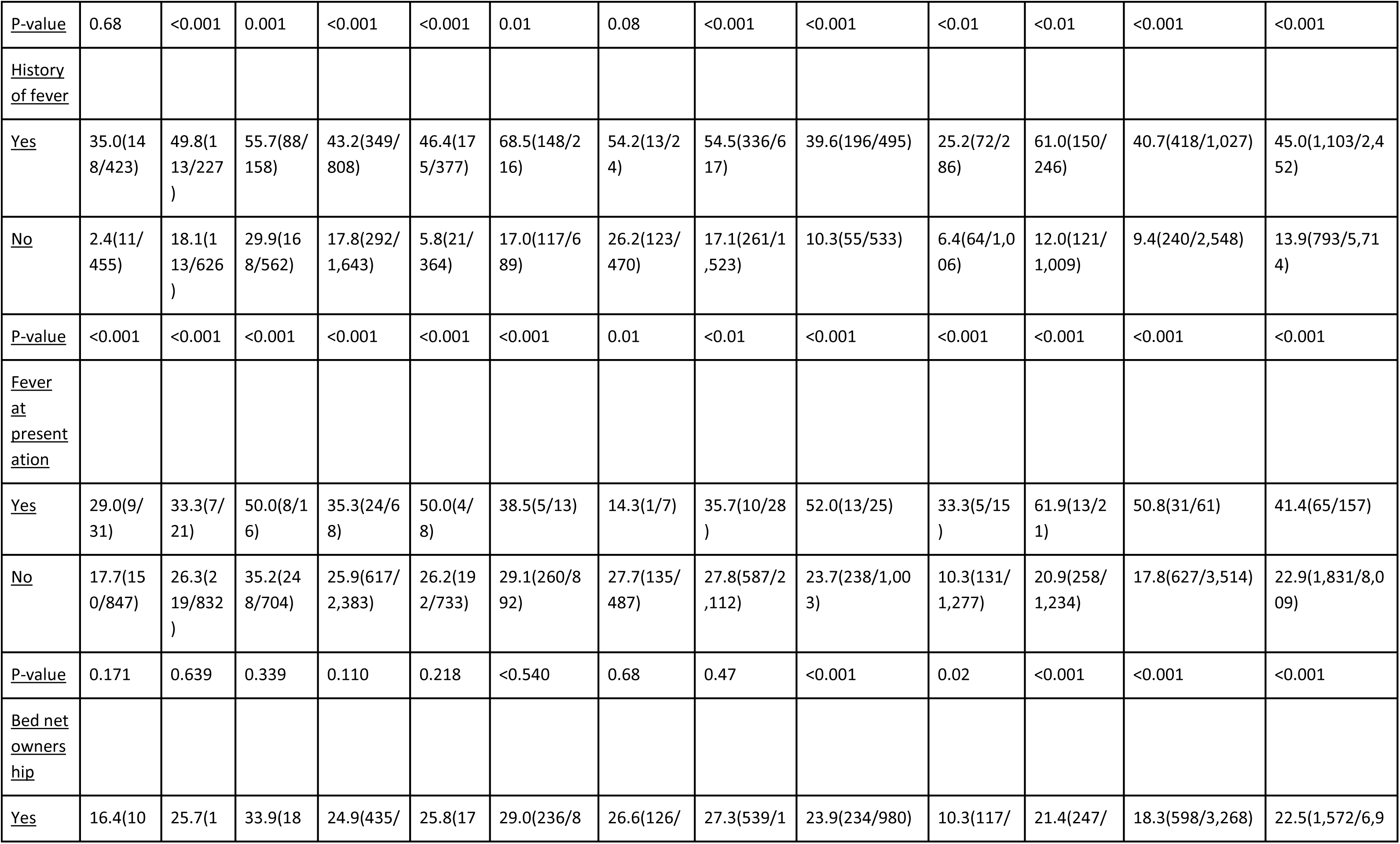
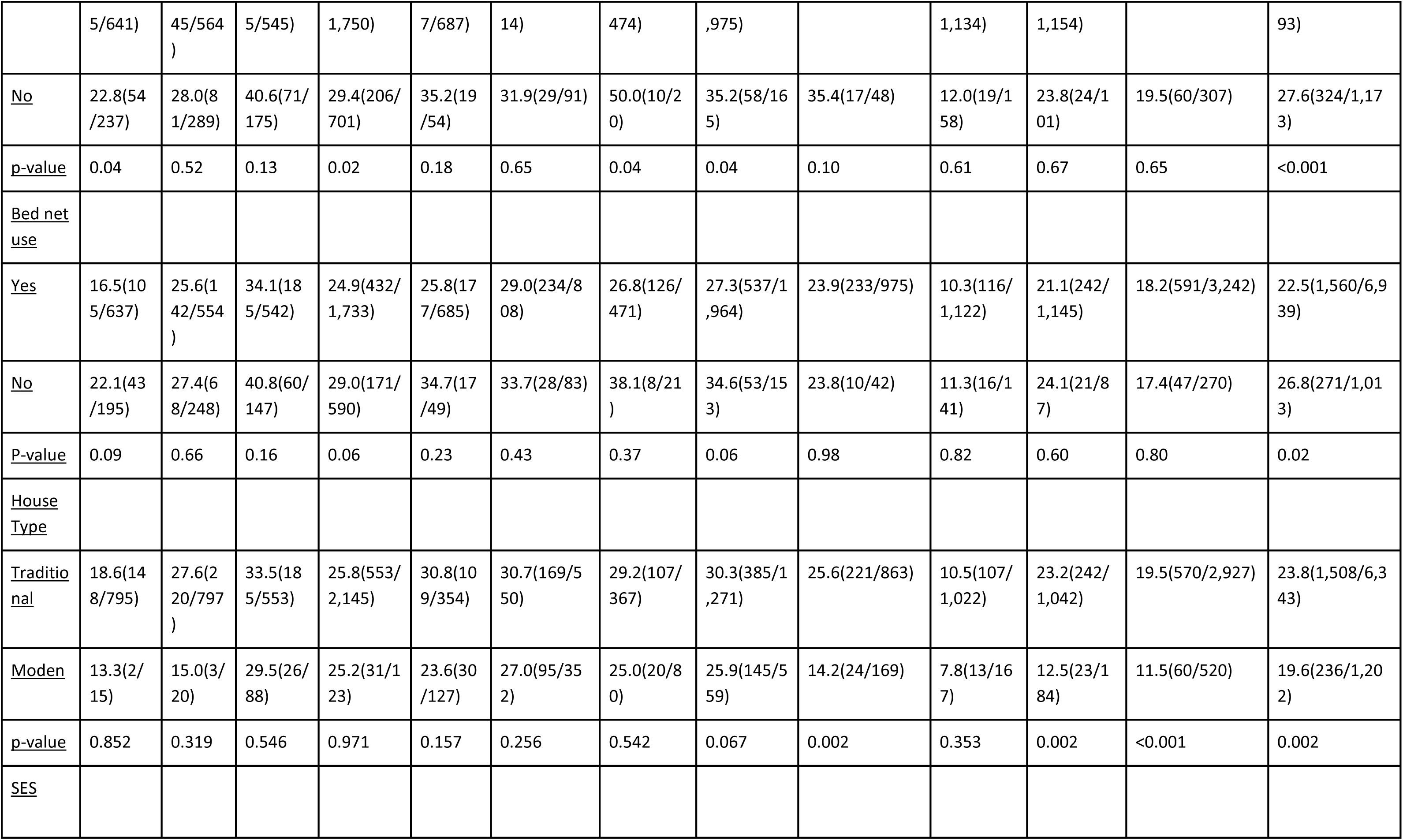
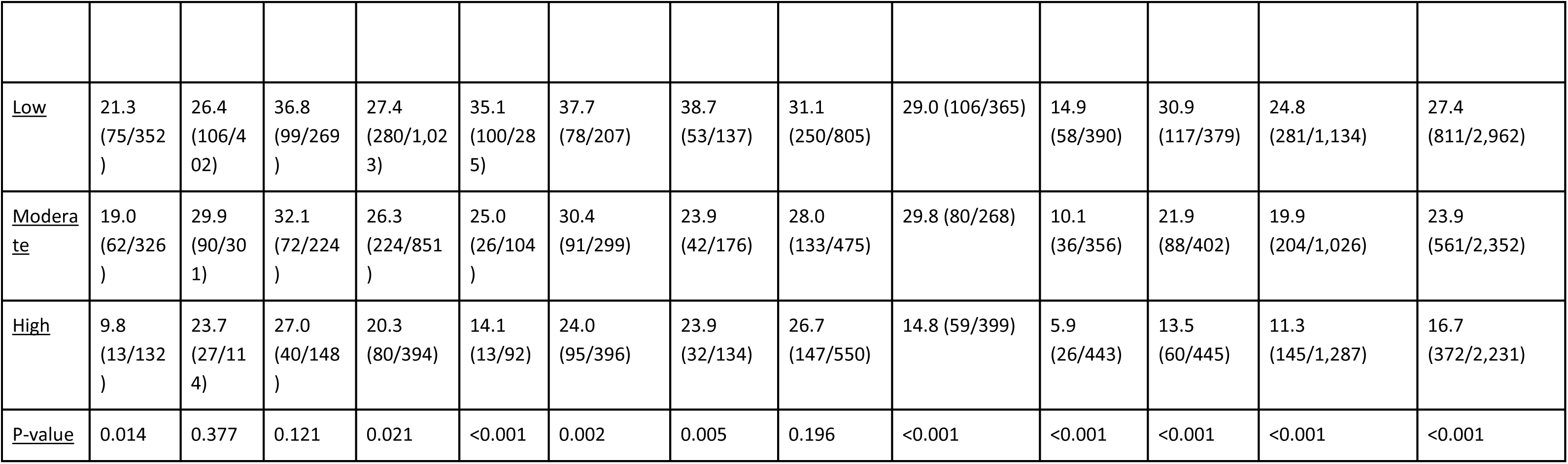
Trends of malaria prevalence (%) in the three regions of Kigoma, Ruvuma and Tanga from 2021 to 2023.

**Supplementary Table 2:**
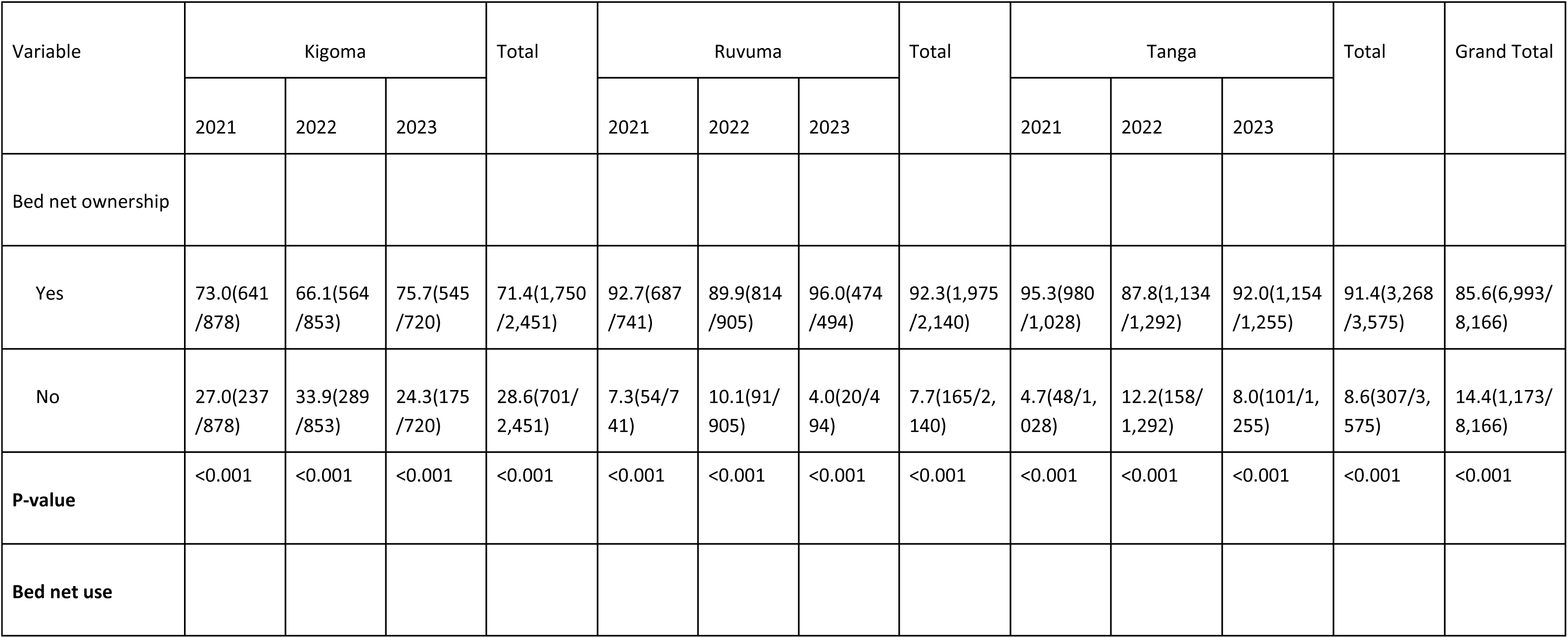
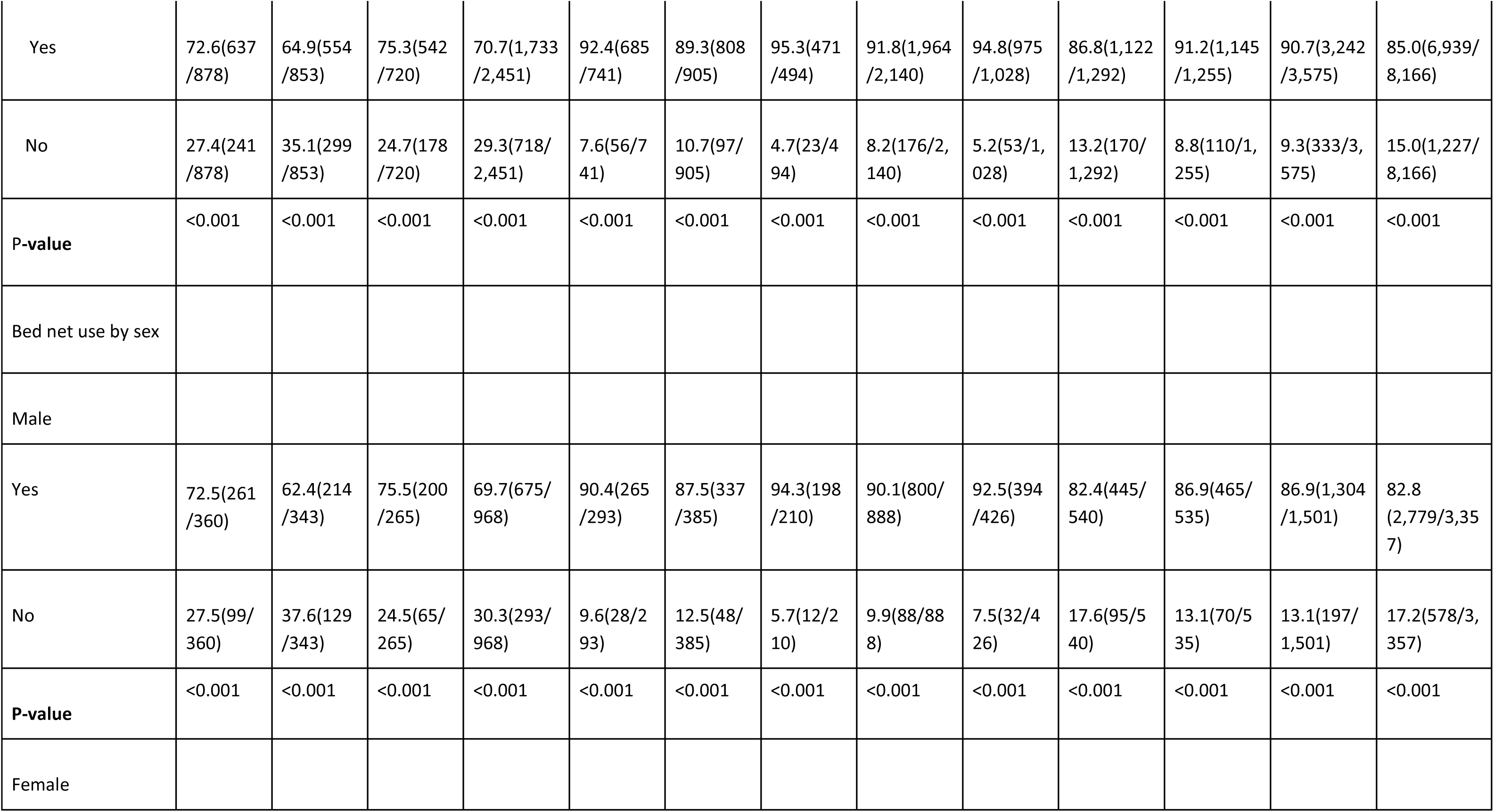
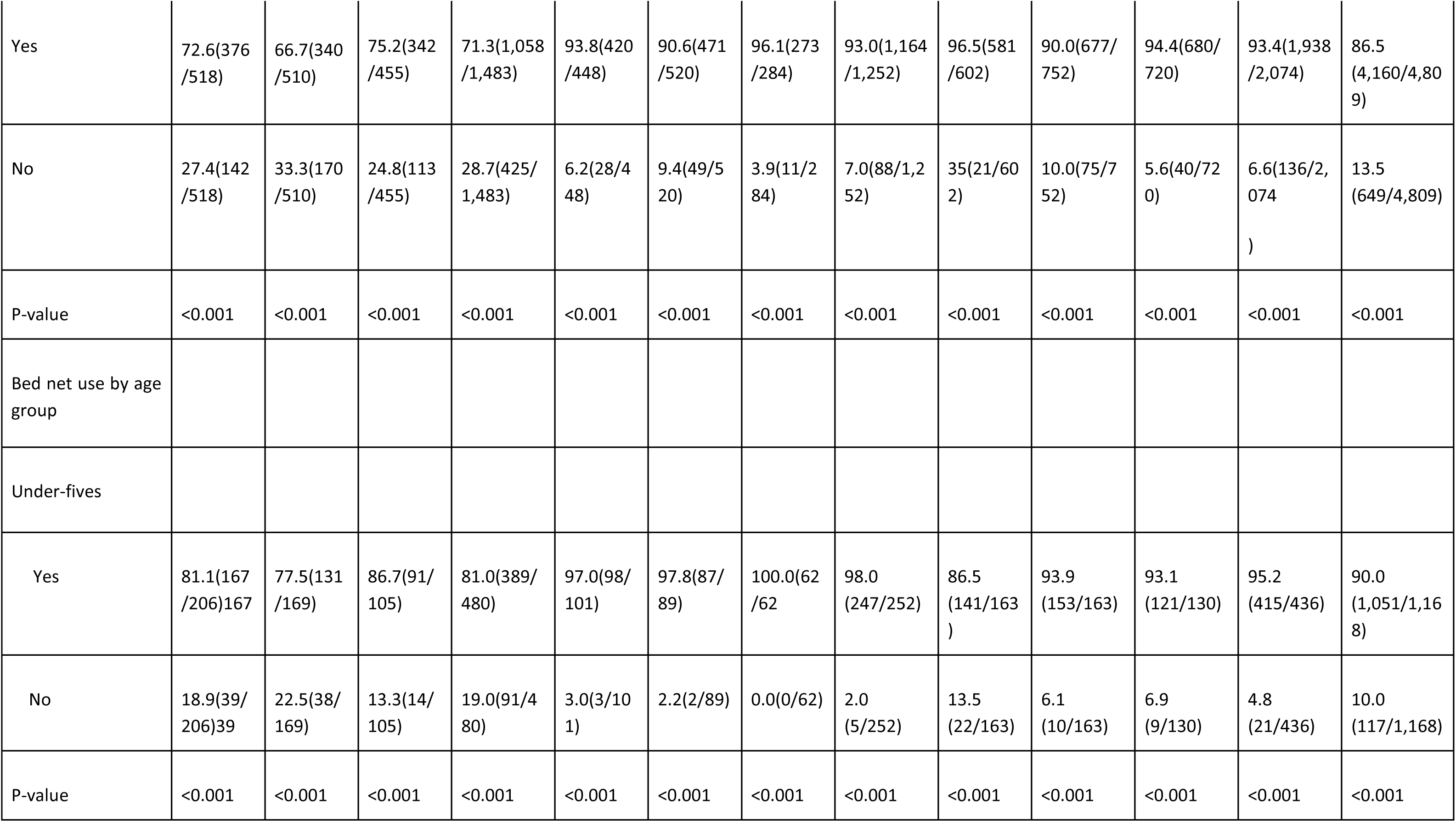
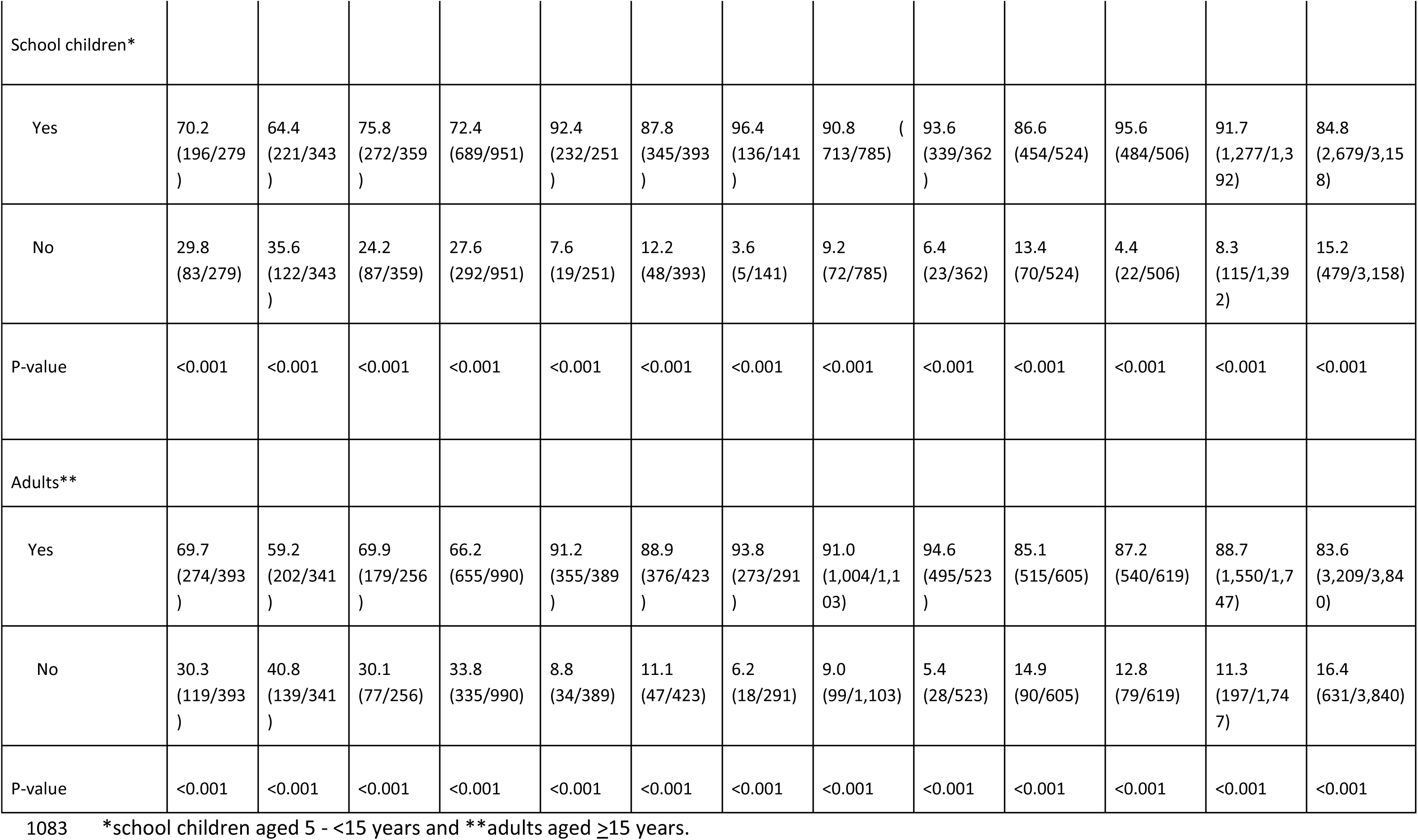
Trends by bed net ownership and use in the three regions from 2021 to 2023.

## Notes

### Competing Interest Statement

The authors have declared no competing interest.

### Funding Statement

This work was supported in full by the Bill and Melinda Gates Foundation grant number 002202. Under the grant conditions of the Foundation, a Creative Commons Attribution 4.0 Generic License has already been assigned to the Author Accepted Manuscript version that might arise from this submission.

### Author Declarations

The ethical approval to conduct this study was obtained from the Medical Research Coordinating Committee of the National Institute for Medical Research (NIMR) in Tanzania. Permission to conduct the study in the villages was provided by the Presidents Office, Regional Administration and Local Government Authority, and health authorities of Tanga, Kigoma, and Ruvuma regions, districts, and villages authorities. Informed consent or assent was sought and obtained before conducting the study, from each participant or parents or legal guardians of children. Permission to publish this paper was sought and given by the Director General of the NIMR.

## References

1. Oladipo HJ, Tajudeen YA, Oladunjoye IO, Yusuff SI, Yusuf RO, Oluwaseyi EM, et al. Increasing challenges of malaria control in sub-Saharan Africa: Priorities for public health research and policymakers. Ann Med Surg (Lond). 2022;81:104366.

2. WHO. World Malaria Report 2024: Addressing inequity in the global malaria response. World Health Organization; 2024.

3. Crutcher 1996 Malaria Medical Microbiology University of Texas Medical Branch at Galveston.pdf.

4. Beeson JG, Brown GV. Pathogenesis of Plasmodium falciparum malaria: the roles of parasite adhesion and antigenic variation. Cell Mol Life Sci. 2002;59:258–71.

5. Kantele A, Jokiranta TS. Review of cases with the emerging fifth human malaria parasite, Plasmodium knowlesi. Clin Infect Dis. 2011;52:1356–62.

6. Belachew EB. Immune Response and Evasion Mechanisms of Plasmodium falciparum Parasites. J Immunol Res. 2018;2018:6529681.

7. Malaria [Internet]. 2024 [cited 2024 Jul 15]. Available from: https://www.cdc.gov/dpdx/malaria/index.html

8. Ishengoma DS, Mmbando BP, Segeja MD, Alfrangis M, Lemnge MM, Bygbjerg IC. Declining burden of malaria over two decades in a rural community of Muheza district, north-eastern Tanzania. Malaria. 2013;12:1–12.

9. Liu Q, Yan W, Qin C, Du M, Liu M, Liu J. Millions of excess cases and thousands of excess deaths of malaria occurred globally in 2020 during the COVID-19 pandemic. J Glob Health. 2022;12:05045.

10. Tanzania Demographic and Health Survey and Malaria Indicator Survey 2022: Key Indicators. Ministry of Health; 2023.

11. Moshi FV. Do early antenatal booking predicts maternal services utilization? An analysis of data from the 2015-16 Tanzania HIV and Malaria Indicators Survey. 2020; Available from: https://www.researchsquare.com/article/rs-35722/latest

12. Challe DP, Kamugisha ML, Mmbando BP, Francis F, Chiduo MG, Mandara CI, et al. Pattern of all-causes and cause-specific mortality in an area with progressively declining malaria burden in Korogwe district, north-eastern Tanzania. Malar J. 2018;17:97.

13. Kitojo C, Gutman JR, Chacky F, Kigadye E, Mkude S, Mandike R, et al. Estimating malaria burden among pregnant women using data from antenatal care centres in Tanzania: a population-based study. Lancet Glob Health. 2019;7:e1695–705.

14. Chacky F, Runge M, Rumisha SF, Machafuko P, Chaki P, Massaga JJ, et al. Nationwide school malaria parasitaemia survey in public primary schools, the United Republic of Tanzania. Malar J. 2018;17:452.

15. Ezezika O, El-Bakri Y, Nadarajah A, Barrett K. Implementation of insecticide-treated malaria bed nets in Tanzania: a systematic review. Journal of Global Health Reports. 2022;6:e2022036.

16. Tungu P, Kabula B, Nkya T, Machafuko P, Sambu E, Batengana B, et al. Trends of insecticide resistance monitoring in mainland Tanzania, 2004-2020. Malar J. 2023;22:100.

17. NMCP. NATIONAL GUIDELINES FOR MALARIA DIAGNOSIS, TREATMENT AND PREVENTIVE THERAPIES 2020 [Internet]. Available from: https://www.nmcp.go.tz/storage/app/uploads/public/643/90d/29e/64390d29ef4b0392189644.pdf

18. Morgan AP, Brazeau NF, Ngasala B, Mhamilawa LE, Denton M, Msellem M, et al. Falciparum malaria from coastal Tanzania and Zanzibar remains highly connected despite effective control efforts on the archipelago. Malar J. 2020;19:47.

19. Koffi AA, Camara S, Ahoua Alou LP, Oumbouke WA, Wolie RZ, Tia IZ, et al. Anopheles vector distribution and malaria transmission dynamics in Gbêkê region, central Côte d’Ivoire. Malar J. 2023;22:192.

20. Rubuga FK, Ahmed A, Siddig E, Sera F, Moirano G, Aimable M, et al. Potential impact of climatic factors on malaria in Rwanda between 2012 and 2021: a time-series analysis. Malar J. 2024;23:274.

21. Mboera LEG, Makundi E a., Kitua AY. Uncertainty in malaria control in Tanzania: crossroads and challenges for future interventions. Am J Trop Med Hyg. 2007;77:112–8.

22. Makundi EA, Mboera LEG, Malebo HM, Kitua AY. Priority setting on malaria interventions in Tanzania: strategies and challenges to mitigate against the intolerable burden. Am J Trop Med Hyg. 2007;77:106–11.

23. Li J, Docile HJ, Fisher D, Pronyuk K, Zhao L. Current status of malaria control and elimination in Africa: Epidemiology, diagnosis, treatment, progress and challenges. J Epidemiol Glob Health. 2024;14:561–79.

24. Ishengoma DS, Mandara CI, Bakari C, Fola AA, Madebe RA, Seth MD, et al. Evidence of artemisinin partial resistance in northwestern Tanzania: clinical and molecular markers of resistance. Lancet Infect Dis. 2024;24:1225–33.

25. Rogier E, Battle N, Bakari C, Seth MD, Nace D, Herman C, et al. Plasmodium falciparum pfhrp2 and pfhrp3 gene deletions among patients enrolled at 100 health facilities throughout Tanzania: February to July 2021. Sci Rep. 2024;14:8158.

26. Liheluka EA, Massawe IS, Chiduo MG, Mandara CI, Chacky F, Ndekuka L, et al. Community knowledge, attitude, practices and beliefs associated with persistence of malaria transmission in North-western and Southern regions of Tanzania. Malar J. 2023;22:1–16.

27. Bakari C, Jones S, Subramaniam G, Mandara CI, Chiduo MG, Rumisha S, et al. Community-based surveys for Plasmodium falciparum pfhrp2 and pfhrp3 gene deletions in selected regions of mainland Tanzania. Malar J. 2020;19:1–12.

28. Guyant P, Corbel V, Guérin PJ, Lautissier A, Nosten F, Boyer S, et al. Past and new challenges for malaria control and elimination: the role of operational research for innovation in designing interventions. Malar J. 2015;14:279.

29. Shretta R, Liu J, Cotter C, Cohen J, Dolenz C, Makomva K, et al. Malaria Elimination and Eradication. In: Holmes KK, Bertozzi S, Bloom BR, Jha P, editors. Major Infectious Diseases. Washington (DC): The International Bank for Reconstruction and Development / The World Bank; 2017.

30. Ishengoma DS, Mmbando BP, Mandara CI, Chiduo MG, Francis F, Timiza W, et al. Trends of Plasmodium falciparum prevalence in two communities of Muheza district North-eastern Tanzania: Correlation between parasite prevalence, malaria interventions and rainfall in the context of re-emergence of malaria after two decades of progressive. Malar J. 2018;17:1–10.

31. Mandai SS, Francis F, Challe DP, Seth MD, Madebe RA, Petro DA, et al. High prevalence and risk of malaria among asymptomatic individuals from villages with high prevalence of artemisinin partial resistance in Kyerwa district of Kagera region, north-western Tanzania. Malar J. 2024;23:197.

32. Mitchell CL, Ngasala B, Janko MM, Chacky F, Edwards JK, Pence BW, et al. Evaluating malaria prevalence and land cover across varying transmission intensity in Tanzania using a cross-sectional survey of school-aged children. Malar J. 2022;21:80.

33. Touray AO, Mobegi VA, Wamunyokoli F, Butungi H, Herren JK. Prevalence of asymptomatic P. falciparum gametocyte carriage among school children in Mbita, Western Kenya and assessment of the association between gametocyte density, multiplicity of infection and mosquito infection prevalence. Wellcome Open Res. 2020;5:259.

34. Juliano JJ, Giesbrecht DJ, Simkin A, Fola AA, Lyimo BM, Pereus D, et al. Country wide surveillance reveals prevalent artemisinin partial resistance mutations with evidence for multiple origins and expansion of high level sulfadoxine-pyrimethamine resistance mutations in northwest Tanzania. medRxiv [Internet]. 2023; Available from: 10.1101/2023.11.07.23298207

35. Popkin Hall ZR, Seth MD, Madebe RA, Budodo R, Bakari C, Francis F, et al. Malaria species positivity rates among symptomatic individuals across regions of differing transmission intensities in Mainland Tanzania. J Infect Dis [Internet]. 2023; Available from: 10.1093/infdis/jiad522

36. Popkin Hall ZR, Seth MD, Madebe RA, Budodo R, Bakari C, Francis F, et al. Malaria species prevalence among asymptomatic individuals in four regions of Mainland Tanzania. medRxiv [Internet]. 2023; Available from: 10.1101/2023.12.28.23300584

37. NMCP. NATIONAL MALARIA STRATEGIC PLAN 2021-2025 TRANSITIONING TO MALARIA ELIMINATION IN PHASES [Internet]. 2020. Available from: http://api-hidl.afya.go.tz/uploads/library-documents/1641210939-jH9mKCtz.pdf

38. Ishengoma DS, Francis F, Mmbando BP, Lusingu JPA, Magistrado P, Alifrangis M, et al. Accuracy of malaria rapid diagnostic tests in community studies and their impact on treatment of malaria in an area with declining malaria burden in north-eastern Tanzania. Malar J. 2011;10:176.

39. Ishengoma DS, Mmbando BP, Segeja MD, Alifrangis M, Lemnge MM. Declining burden of malaria over two decades in a rural community of Muheza district,. Malar J. 2013;338:1–12.

40. Francis F, Ishengoma DS, Mmbando BP, Rutta ASM, Malecela MN, Mayala B, et al. Deployment and use of mobile phone technology for real-time reporting of fever cases and malaria treatment failure in areas of declining malaria transmission in Muheza district north-eastern Tanzania. Malar J. 2017;16:308.

41. Chacha GA, Francis F, Mandai SS, Seth MD, Madebe RA, Challe DP, et al. Prevalence and drivers of malaria infection among asymptomatic and symptomatic community members in five regions with varying transmission intensity in mainland Tanzania. Parasit Vectors [Internet]. 2025;18. Available from: 10.1186/s13071-024-06639-1

42. Kamugisha ML, Mmbando BP, Francis F, Ishengoma DS, Challe DP, Lemnge MM. Establishing and implementing Demographic Surveillance System as a tool for monitoring health interventions in Korogwe District, northastern Tanzania. Tanzan J Health Res. 2011;13:57–67.

43. Challe 2024 Prevalence and risk factors associated with malaria infections at micro-geographic level in three villages of Muheza district. north-eastern Tanzania;

44. Chacha GA, Francis F, Mandai SS, Seth MD, Madebe RA, Challe DP, et al. Prevalence and drivers of malaria infections among asymptomatic individuals from selected communities in five regions of Mainland Tanzania with varying transmission intensities [Internet]. bioRxiv. 2024. Available from: 10.1101/2024.06.05.24308481

45. Kayendeke M, Nabirye C, Nayiga S, Westercamp N, Gonahasa S, Katureebe A, et al. House modifications as a malaria control tool: how does local context shape participants’ experience and interpretation in Uganda? Malar J. 2023;22:244.

46. Pinder M, Bradley J, Jawara M, Affara M, Conteh L, Correa S, et al. Improved housing versus usual practice for additional protection against clinical malaria in The Gambia (RooPfs): a household-randomised controlled trial. Lancet Planet Health. 2021;5:e220–9.

47. Popkin-Hall ZR, Seth MD, Madebe RA, Budodo R, Bakari C, Francis F, et al. Prevalence of non-falciparum malaria infections among asymptomatic individuals in four regions of Mainland Tanzania. Parasit Vectors. 2024;17:153.

48. MoH 2017 Standard treatment guidelines & National Essential Medicines list Tanzania Mainland.pdf.

49. Nawa M, Mupeyo-Mudala C, Banda-Tembo S, Adetokunboh O. The effects of modern housing on malaria transmission in different endemic zones: a systematic review and meta-analysis. Malar J. 2024;23:235.

50. Fox T, Furnival-Adams J, Chaplin M, Napier M, Olanga EA. House modifications for preventing malaria. Cochrane Database Syst Rev. 2022;10:CD013398.

51. Mmbando BP, Kamugisha ML, Lusingu JP, Francis F, Ishengoma DS, Theander TG, et al. Spatial variation and socio-economic determinants of Plasmodium falciparum infection in northeastern Tanzania. Malar J. 2011;10:1–9.

52. Rumisha SF, Shayo EH, Mboera LEG. Spatio-temporal prevalence of malaria and anaemia in relation to agro-ecosystems in Mvomero district, Tanzania. Malar J. 2019;18:228.

53. Susanna D, Pratiwi D. Current status of insecticide resistance in malaria vectors in the Asian countries: a systematic review. F1000Res. 2021;10:200.

54. Suh PF, Elanga-Ndille E, Tchouakui M, Sandeu MM, Tagne D, Wondji C, et al. Impact of insecticide resistance on malaria vector competence: a literature review. Malar J. 2023;22:19.

55. WHO_CDS_RBM_ Malaria early warning systems _2001.32.pdf.

56. Lindblade KA, Walker ED, Onapa AW, Katungu J, Wilson ML. Land use change alters malaria transmission parameters by modifying temperature in a highland area of Uganda. Trop Med Int Health. 2000;5:263–74.

57. Nabatanzi M, Ntono V, Kamulegeya J, Kwesiga B, Bulage L, Lubwama B, et al. Malaria outbreak facilitated by increased mosquito breeding sites near houses and cessation of indoor residual spraying, Kole district, Uganda, January-June 2019. BMC Public Health. 2022;22:1898.

58. Ishengoma DS, Mandara CI, Francis F, Talundzic E, Lucchi NW, Ngasala B, et al. Efficacy and safety of artemether-lumefantrine for the treatment of uncomplicated malaria and prevalence of Pfk13 and Pfmdr1 polymorphisms after a decade of using artemisinin-based combination therapy in mainland Tanzania. Malar J. 2019;18:88.

59. Beke OA-H, Assi S-B, Kokrasset APH, Dibo KJD, Tanoh MA, Danho M, et al. Implication of agricultural practices in the micro-geographic heterogeneity of malaria transmission in Bouna, Côte d’Ivoire. Malar J. 2023;22:313.

60. Sharma RK, Singh MP, Saha KB, Bharti PK, Jain V, Singh PP, et al. Socio-economic & household risk factors of malaria in tribal areas of Madhya Pradesh, central India. Indian J Med Res. 2015;141:567–75.

61. Munajat MB, Rahim MAFA, Wahid W, Seri Rakna MIM, Divis PCS, Chuangchaiya S, et al. Perceptions and prevention practices on malaria among the indigenous Orang Asli community in Kelantan, Peninsular Malaysia. Malar J. 2021;20:202.

62. Scott J, Kanyangarara M, Nhama A, Macete E, Moss WJ, Saute F. Factors associated with use of insecticide-treated net for malaria prevention in Manica District, Mozambique: a community-based cross-sectional survey. Malar J. 2021;20:1–9.

63. Nzobo BJ, Ngasala BE, Kihamia CM. Prevalence of asymptomatic malaria infection and use of different malaria control measures among primary school children in Morogoro Municipality, Tanzania. Malar J. 2015;14:1–7.

64. Chilanga E, Collin-Vézina D, MacIntosh H, Mitchell C, Cherney K. Prevalence and determinants of malaria infection among children of local farmers in Central Malawi. Malar J. 2020;19:308.

65. Habyarimana F, Ramroop S. Prevalence and risk factors associated with malaria among children aged six months to 14 years old in Rwanda: Evidence from 2017 Rwanda Malaria Indicator Survey. Int J Environ Res Public Health. 2020;17:7975.

66. Barua P, Beeson JG, Maleta K, Ashorn P, Rogerson SJ. The impact of early life exposure to Plasmodium falciparum on the development of naturally acquired immunity to malaria in young Malawian children. Malar J. 2019;18:11.

67. Isiko I, Nyegenye S, Bett DK, Asingwire JM, Okoro LN, Emeribe NA, et al. Factors associated with the risk of malaria among children: analysis of 2021 Nigeria Malaria Indicator Survey. Malar J. 2024;23:109.

68. Clarke SE, Rouhani S, Diarra S, Saye R, Bamadio M, Jones R, et al. Impact of a malaria intervention package in schools on Plasmodium infection, anaemia and cognitive function in schoolchildren in Mali: a pragmatic cluster-randomised trial. BMJ Glob Health. 2017;2:e000182.

69. Kihwele F, Gavana T, Makungu C, Msuya HM, Mlacha YP, Govella NJ, et al. Exploring activities and behaviours potentially increases school-age children’s vulnerability to malaria infections in south-eastern Tanzania. Malar J. 2023;22:1–11.

70. Akello AR, Byagamy JP, Etajak S, Okadhi CS, Yeka A. Factors influencing consistent use of bed nets for the control of malaria among children under 5 years in Soroti District, North Eastern Uganda. Malar J. 2022;21:1–13.

71. González-Sanz M, Berzosa P, Norman FF. Updates on Malaria Epidemiology and Prevention Strategies. Curr Infect Dis Rep. 2023;1–9.

72. Onyinyechi OM, Mohd Nazan AIN, Ismail S. Effectiveness of health education interventions to improve malaria knowledge and insecticide-treated nets usage among populations of sub-Saharan Africa: systematic review and meta-analysis. Front Public Health. 2023;11:1217052.

73. Aponte JJ, Menendez C, Schellenberg D, Kahigwa E, Mshinda H, Vountasou P, et al. Age interactions in the development of naturally acquired immunity to Plasmodium falciparum and its clinical presentation. PLoS Med. 2007;4:e242.

74. Abbas F, Kigadye E, Mohamed F, Khamis M, Mbaraka J, Serbantez N, et al. Socio-demographic trends in malaria knowledge and implications for behaviour change interventions in Zanzibar. Malar J. 2023;22:39.

75. Rahmani AA, Susanna D, Febrian T. The relationship between climate change and malaria in South-East Asia: A systematic review of the evidence. F1000Res. 2022;11:1555.

76. Abdalal SA, Yukich J, Andrinopoulos K, Alghanmi M, Wakid MH, Zawawi A, et al. Livelihood activities, human mobility, and risk of malaria infection in elimination settings: a case-control study. Malar J. 2023;22:53.

77. Lwenge M, Govule P, Katongole SP, Dako-Gyeke P. Malaria treatment health seeking behaviors among international students at the University of Ghana Legon. PLoS One. 2023;18:e0276412.

78. Tadesse 2021 Malaria prevention and treatment in migrant agricultural workers in Dangur district. social and behavioural aspects. Ethiopia;

79. Abdalal 2023 Livelihood activities, human mobility, and risk of malaria infection in elimination settings: a case-control study.

80. Mandefro A, Tadele G, Mekonen B, Golassa L. Analysing the six-year malaria trends at Metehara Health Centre in Central Ethiopia: the impact of resurgence on the 2030 elimination goals. Malar J. 2024;23:32.

81. Okiring J, Epstein A, Namuganga JF, Kamya EV, Nabende I, Nassali M, et al. Gender difference in the incidence of malaria diagnosed at public health facilities in Uganda. Malar J. 2022;21:22.

82. Moshi 2028 Outdoor malaria transmission risks and social life: a qualitative study in South-Eastern Tanzania.

83. Latunji OO, Akinyemi OO. FACTORS INFLUENCING HEALTH-SEEKING BEHAVIOUR AMONG CIVIL SERVANTS IN IBADAN, NIGERIA. Ann Ib Postgrad Med. 2018;16:52–60.

84. Alhassan Y, Dwomoh D, Amuasi SA, Nonvignon J, Bonful H, Tetteh M, et al. Impact of insecticide-treated nets and indoor residual spraying on self-reported malaria prevalence among women of reproductive age in Ghana: implication for malaria control and elimination. Malar J. 2022;21:120.

85. Taremwa IM, Ashaba S, Kyarisiima R, Ayebazibwe C, Ninsiima R, Mattison C. Treatment-seeking and uptake of malaria prevention strategies among pregnant women and caregivers of children under-five years during COVID-19 pandemic in rural communities in South West Uganda: a qualitative study. BMC Public Health. 2022;22:373.

86. Cervantes-Candelas LA, Aguilar-Castro J, Buendía-González FO, Fernández-Rivera O, Nolasco-Pérez T de J, López-Padilla MS, et al. 17β-Estradiol Is Involved in the Sexual Dimorphism of the Immune Response to Malaria. Front Endocrinol. 2021;12:643851.

87. Simmons RA, Mboera L, Miranda ML, Morris A, Stresman G, Turner EL, et al. A longitudinal cohort study of malaria exposure and changing serostatus in a malaria endemic area of rural Tanzania. Malar J [Internet]. 2017;16. Available from: 10.1186/s12936-017-1945-2

88. Huang F, Takala-Harrison S, Liu H, Xu J-W, Yang H-L, Adams M, et al. Prevalence of Clinical and Subclinical Plasmodium falciparum and Plasmodium vivax Malaria in Two Remote Rural Communities on the Myanmar-China Border. Am J Trop Med Hyg. 2017;97:1524–31.

89. Melese Y, Alemu M, Yimer M, Tegegne B, Tadele T. Asymptomatic Malaria in Households and Neighbors of Laboratory Confirmed Cases in Raya Kobo District, Northeast Ethiopia. Ethiop J Health Sci. 2022;32:623–30.

90. Kamau A, Mtanje G, Mataza C, Mwambingu G, Mturi N, Mohammed S, et al. Malaria infection, disease and mortality among children and adults on the coast of Kenya. Malar J. 2020;19:210.

91. Houben CH, Fleischmann H, Gückel M. Malaria prevalence in north-eastern Nigeria: a cross-sectional study. Asian Pac J Trop Med. 2013;6:865–8.

92. Paasi G, Ndila CM, Okiror W, Namayanja C, Okalebo BP, Abongo G, et al. Characterising childhood Blackwater Fever and its clinical care at two tertiary hospitals in Eastern Uganda. 2021; Available from: https://www.researchsquare.com/article/rs-545832/v1

93. Emiru T, Getachew D, Murphy M, Sedda L, Ejigu LA, Bulto MG, et al. Evidence for a role of Anopheles stephensi in the spread of drug- and diagnosis-resistant malaria in Africa. Nat Med. 2023;29:3203–11.

94. Madut DB, Rubach MP, Bonnewell JP, Cutting ER, Carugati M, Kalengo N, et al. Trends in fever case management for febrile inpatients in a low malaria incidence setting of Tanzania. Trop Med Int Health. 2021;26:1668–76.

95. Marinkovic M, Diez-Silva M, Pantic I, Fredberg JJ, Suresh S, Butler JP. Febrile temperature leads to significant stiffening of Plasmodium falciparum parasitized erythrocytes. Am J Physiol Cell Physiol. 2009;296:C59–64.

96. Babalola S, Adedokun ST, McCartney-Melstad A, Okoh M, Asa S, Tweedie I, et al. Factors associated with caregivers’ consistency of use of bed nets in Nigeria: a multilevel multinomial analysis of survey data. Malar J. 2018;17:280.

97. Zerdo Z, Bastiaens H, Anthierens S, Massebo F, Masne M, Biresaw G, et al. Effect of malaria prevention education on bed net utilization, incidence of malaria and treatment seeking among school-aged children in Southern Ethiopia; cluster randomized controlled trial. BMC Infect Dis. 2023;23:486.

98. Haq IU, Mehmood Z, Khan GA, Kainat B, Ahmed B, Shah J, et al. Modeling the effect of climatic conditions and topography on malaria incidence using Poisson regression: a Retrospective study in Bannu, Khyber Pakhtunkhwa, Pakistan. Front Microbiol. 2023;14:1303087.

99. Nankabirwa JI, Gonahasa S, Katureebe A, Mutungi P, Nassali M, Kamya MR, et al. The Uganda Housing Modification Study – Association between housing characteristics and malaria burden in a moderate to high transmission setting in Uganda [Internet]. Research Square. 2024. Available from: 10.21203/rs.3.rs-4319094/v1

100. Bofu RM, Santos EM, Msugupakulya BJ, Kahamba NF, Swilla JD, Njalambaha R, et al. The needs and opportunities for housing improvement for malaria control in southern. Malar J. 2023;22:1–15.

101. Zong L, Ngarukiyimana JP, Yang Y, Yim SHL, Zhou Y, Wang M, et al. Malaria transmission risk is projected to increase in the highlands of Western and Northern Rwanda. Commun Earth Environ [Internet]. 2024;5. Available from: 10.1038/s43247-024-01717-9

102. Castro MC. Malaria transmission and prospects for malaria eradication: The role of the environment. Cold Spring Harb Perspect Med. 2017;7:a025601.

103. Baragatti M, Fournet F, Henry M-C, Assi S, Ouedraogo H, Rogier C, et al. Social and environmental malaria risk factors in urban areas of Ouagadougou, Burkina Faso. Malar J. 2009;8:13.

104. Budodo 2024 Performance of rapid diagnostic tests, microscopy, and qPCR for detection of parasites among community members with or without symptoms of malaria in villages with high levels of artemisinin partial. North-western Tanzania;

